# Increased mortality in community-tested cases of SARS-CoV-2 lineage B.1.1.7

**DOI:** 10.1101/2021.02.01.21250959

**Authors:** Nicholas G. Davies, Christopher I. Jarvis, CMMID COVID-19 Working Group, W. John Edmunds, Nicholas P. Jewell, Karla Diaz-Ordaz, Ruth H. Keogh

## Abstract

SARS-CoV-2 lineage B.1.1.7, a variant first detected in the United Kingdom in September 2020^1^, has spread to multiple countries worldwide. Several studies have established that B.1.1.7 is more transmissible than preexisting variants, but have not identified whether it leads to any change in disease severity^2^. We analyse a dataset linking 2,245,263 positive SARS-CoV-2 community tests and 17,452 COVID-19 deaths in England from 1 September 2020 to 14 February 2021. For 1,146,534 (51%) of these tests, the presence or absence of B.1.1.7 can be identified because of mutations in this lineage preventing PCR amplification of the spike gene target (S gene target failure, SGTF^1^). Based on 4,945 deaths with known SGTF status, we estimate that the hazard of death associated with SGTF is 55% (95% CI 39–72%) higher after adjustment for age, sex, ethnicity, deprivation, care home residence, local authority of residence and test date. This corresponds to the absolute risk of death for a 55–69-year-old male increasing from 0.6% to 0.9% (95% CI 0.8–1.0%) within 28 days after a positive test in the community. Correcting for misclassification of SGTF and missingness in SGTF status, we estimate a 61% (42–82%) higher hazard of death associated with B.1.1.7. Our analysis suggests that B.1.1.7 is not only more transmissible than preexisting SARS-CoV-2 variants, but may also cause more severe illness.

---

Most community SARS-CoV-2 PCR tests in England are processed by one of six national “Lighthouse” laboratories. Among the mutations carried by lineage B.1.1.7—also known as Variant of Concern (VOC) 202012/01—is a 6-nucleotide deletion that prevents amplification of the S gene target by the commercial PCR assay currently used in three of the Lighthouse labs^1^. By linking individual records of positive community tests with and without S gene target failure (SGTF) to a comprehensive line list of COVID-19 deaths in England, we estimate the relative hazard of death associated with B.1.1.7 infection. We define confirmed SGTF as a compatible PCR result with cycle threshold (Ct) < 30 for ORF1ab, Ct < 30 for N, and no detectable S (Ct > 40); confirmed non-SGTF as any compatible PCR result with Ct < 30 for each of ORF1ab, N, and S; and an inconclusive (missing) result as any other positive test, including tests processed by a laboratory incapable of assessing SGTF.

## Characteristics of the study population

The study sample (**Extended Data Table 1**) comprises 2,245,263 individuals who had a positive community (“Pillar 2”) test between 1 November 2020 and 14 February 2021. Just over half of those tested (1,146,534, 51.1%) had a conclusive SGTF reading and, of these, 58.8% had SGTF. Females comprised 53.6% of the total sample; 44.3% were aged 1–34 years, 34.4% aged 35–54, 15.1% aged 55–69, 4.3% aged 70–84 and 1.9% aged 85 or older. The majority of individuals (93.7%) lived in residential accommodation (defined as a house, flat, sheltered accommodation, or house in multiple occupancy), with 3.1% living in a care or nursing home. Based on self-identified ethnicity, 74.0% were White, 13.6% were Asian, 4.6% were Black and 7.8% were of other, mixed or unknown ethnicity. All seven NHS England regions are represented, with the London region contributing 22.5% of tests and the South West 5.9%. The first three weeks of the study period (1–21 November) contributed 15.5% of the total tests, and the final three weeks (24 January–14 February) 12.8%. The period between 3–23 January contributed 31.6% of tests.

In those with SGTF status measured, SGTF prevalence was similar in males and females but lower in the older age groups: 59.0% in 1–34-year-olds compared with 55.4% in those aged 85 and older. In keeping with these age patterns, SGTF prevalence was lower in individuals living in a care or nursing home (54.3%) than those in residential accommodation (58.8%). SGTF prevalence by self-identified ethnicity was 58.0% in the White group, 57.6% in the Asian group, 69.6% in the Black group, and 64.8% in the other, mixed, or unknown ethnicity group. SGTF prevalence was lowest in the most deprived index of multiple deprivation^3^ (IMD) quintile (53.9%). The highest prevalences of SGTF over the study period were observed in the East of England (77.5%), South East (77.3%) and London (75.4%) regions, and prevalence of SGTF was lowest in the North East and Yorkshire region (41.2%). The prevalence of SGTF increased steeply over time (**Fig. 1a**), from 5.8% during 1–21 November 2020 to 94.3% during 24 January–14 February 2021.

**Fig. 1.**
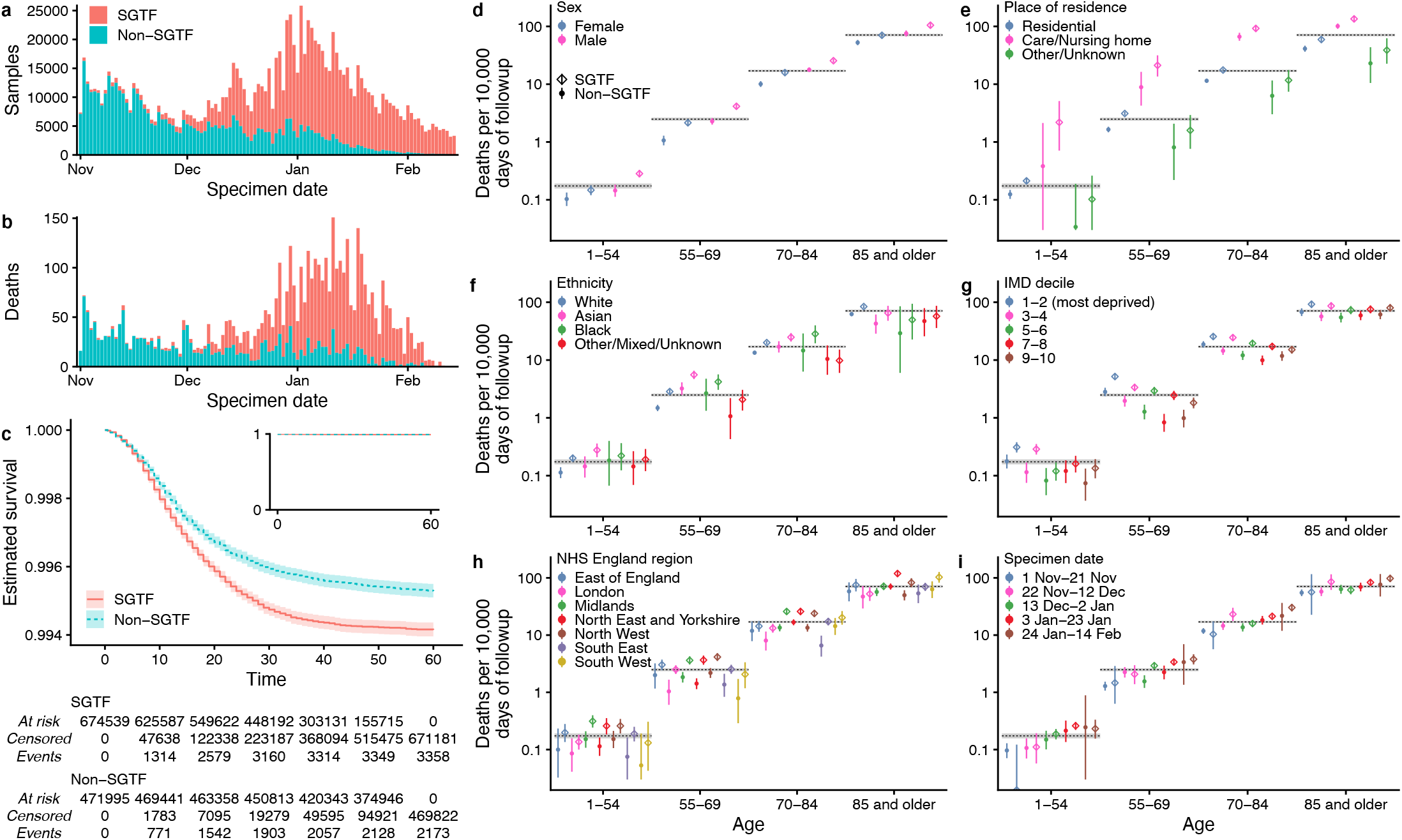
Descriptive analyses. **a** The number of samples with and without SGTF by day from 1 November 2020 to 14 February 2021, the period covered by our main analysis. **b** Number of deaths within 28 days of positive test by specimen date included in the analysis. **c** Kaplan-Meier plot showing survival (95% confidence intervals) among individuals tested in the community in England with and without SGTF, in the subset with SGTF measured. Inset shows the full *y*–axis range. **d–i** Crude death rates (point estimates and 95% CIs) among SGTF versus non-SGTF cases (in the subset with SGTF measured, *n* = 1,146,534) for deaths within 28 days of positive test stratified by broad age groups and (**d**) sex, (**e**) place of residence, (**f**) ethnicity, (**g**) index of multiple deprivation, (**h**) NHS England region, and (**i**) specimen date. Horizontal bars show the overall crude death rates (95% CIs) by age group irrespective of SGTF status.

Missing SGTF status was strongly associated with age and place of residence. The proportion with SGTF status missing was similar in age groups 1–34 (48.3%), 35–54 (47.8%) and 55–69 (48.2%), and then rose to 54.4% in the 70–84 age group and to 77.7% in the 85 and older age group. SGTF status was missing in 87.9% of tests for individuals living in a care or nursing home, compared to 47.4% of tests among individuals in residential accommodation. This is partly due to more extensive use of lateral flow immunoassay tests in care homes, which do not yield an SGTF reading. Missingness in SGTF status also differed substantially by NHS England region, ranging from 21.2% in the North West to 71.1% in the South West, which is largely explained by proximity to a Lighthouse lab capable of producing an SGTF reading (**Extended Data Fig. 1**). Missingness also depended on specimen date, with the percentage missing being lower for the earlier specimen dates and highest (54.4%) in the 21-day period that contributed the most tests (3–23 January). There were also minor differences in missingness by ethnicity and IMD. Of the 48.9% of tests with missing SGTF status, 5.1% were inconclusive due to high Ct values and the remaining 43.8% were not assessed for SGTF.

19,615 people in the study sample are known to have died (0.87% of 2,245,263). Crude death rates were substantially higher in the elderly and in those living in a care or nursing home (**Supplementary Table 1**). The standard definition of a COVID-19 death in England is any death occurring within 28 days of an individual’s first positive SARS-CoV-2 test; 17,452 of the observed deaths (89.0%) met this criterion (**Fig. 1b**). Among those with known SGTF status, the crude COVID-19 death rate was 1.86 deaths per 10,000 person-days of follow-up in the SGTF group, versus 1.42 deaths per 10,000 person-days in the non-SGTF group (**Fig. 1c**; **Extended Data Table 2**). Stratifying by broad age groups and by sex, place of residence, ethnicity, IMD, region, and specimen date, death rates within 28 days of a positive SARS-CoV-2 test were higher among SGTF than non-SGTF cases in 98 of the 104 strata assessed (94%; **Figs. 1d–i**; see also **Supplementary Table 2**).

## Cox regression analyses

To estimate the effect of SGTF on mortality while controlling for observed confounding (**Extended Data Fig. 2**), we fitted a series of Cox proportional hazards models^4^ to the data. We stratified the analysis by lower tier local authority (LTLA) and specimen date to control for geographical and temporal differences in the baseline hazard—for example, due to changes in hospital pressure during the study period—and used spline terms for age and IMD and fixed effects for sex, ethnicity, and residence type. All models were fitted twice, once using complete cases only, i.e. by simply excluding individuals with missing SGTF status, and once using inverse probability weighting (IPW), i.e. accounting for missingness by upweighting individuals whose characteristics—age, sex, IMD, ethnicity, residence type, NHS England region of residence and sampling week—are underrepresented among complete cases. This analysis assumes that, holding these characteristics constant, whether an individual dies is independent of missingness in SGTF status^5^.

For the complete-cases analysis, the estimated hazard ratio for SGTF was 1.55 (95% CI 1.39– 1.72), indicating that the hazard of death in the 28 days following a positive test is 55% (39– 72%) higher for SGTF than for non-SGTF cases.

To assess the model assumption of proportional hazards, we added an interaction term between SGTF and time since positive test. There was strong evidence of non-proportionality of hazards (likelihood ratio test 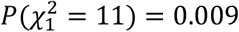; **Fig. 2a**; **Extended Data Fig. 3**), with the estimated time-varying hazard ratio increasing over time: 1.14 (0.92–1.40) one day after the positive test, 1.58 (1.42–1.75) on day 14, and 2.24 (1.75–2.87) on day 28. Adding higher-order functions of time into the interaction terms did not significantly improve model fit (likelihood ratio test 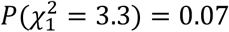). We found no evidence that the effect of SGTF varied by age group (likelihood ratio test 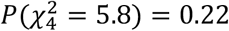), sex 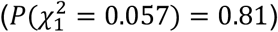, IMD 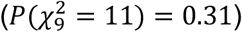, ethnicity 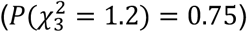, or residence type 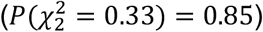. We note, however, that the relatively small number of deaths among 1–34-year-olds over the study period (44 deaths) does not permit robust assessment of the impact of SGTF in this age group. Other time-covariate interactions suggested that the delay from positive test to death was slightly shorter among females, care home residents, and the elderly; see **Supplementary Note 1** for more details on models with interaction terms.

**Fig. 2.**
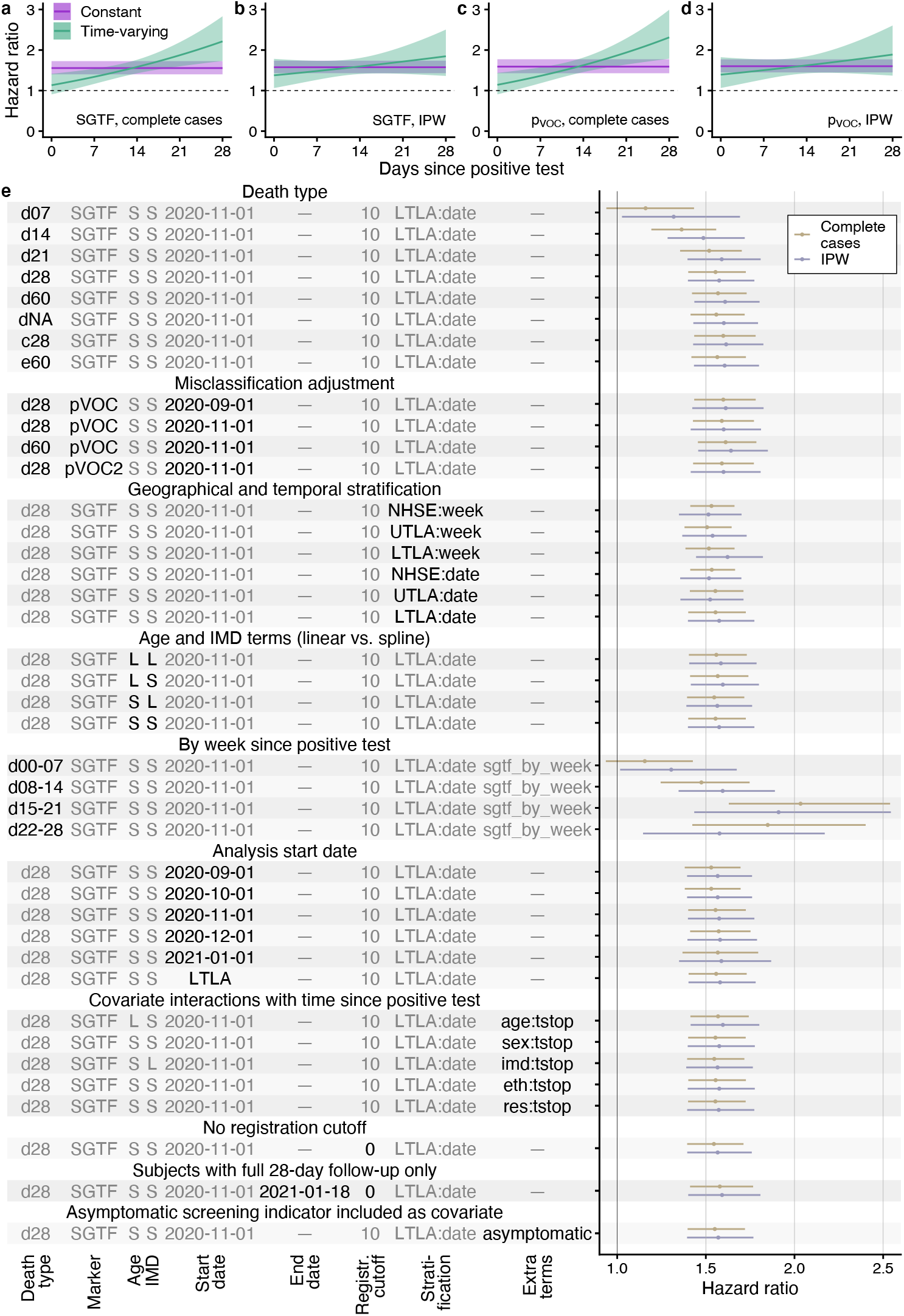
Survival analyses. **a–d** Estimated hazard ratio of death (mean and 95% CIs) within 28 days of positive test for (**a**) SGTF, complete-cases analysis; (**b**) SGTF, IPW analysis; (**c**) *p*_VOC_, complete-cases analysis; and (**d**) *p*_VOC_, IPW analysis, in model stratified by LTLA and specimen date and adjusted for the other covariates. **e** Estimated hazard ratio of death (point estimates and 95% CIs) across each model investigated. Death types are coded as follows: d*X*, all deaths within *X* days of a positive test; c28, death-certificate-confirmed COVID-19 deaths within 28 days; e60, all deaths within 60 days plus all death-certificate-confirmed COVID-19 deaths within any time period. S, spline term (for Age or IMD); L, linear term (for Age or IMD); NHSE, NHS England region (*n* = 7); UTLA, upper-tier local authority (*n* = 150); LTLA, lower-tier local authority (*n* = 316). LTLA start date signifies a start date chosen separately for each LTLA (see Methods).

For IPW analysis, a model to predict missingness is required. We evaluated a series of such models, including a cauchit model, which is a robust alternative to logistic regression suitable for IPW^5^. We selected the cauchit model as it fit well and resulted in less extreme weights than other models (**Extended Data Fig. 4**). The IPW analysis gave similar results to the complete-cases analysis, yielding a hazard ratio of 1.58 (1.40–1.78). Like the complete-cases analysis, the IPW analysis recovered an increasing hazard with time since positive test, but the increase was less marked (**Fig. 2b**) and did not significantly differ from zero (Wald test 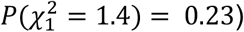.

## Misclassification analysis

Prior to the emergence of B.1.1.7, a number of minor circulating SARS-CoV-2 lineages with spike mutations could also cause SGTF^1^. Our main analyses are restricted to specimens from 1 November 2020 onwards to avoid diluting the measured effect of B.1.1.7 on mortality due to non-B.1.1.7 lineages causing SGTF. As an alternative approach, we undertook a misclassification analysis^6^, modelling the relative frequency of SGTF over time for each NHS England region as a low, time-invariant frequency of non-B.1.1.7 samples with SGTF plus a logistically growing^2^ frequency of B.1.1.7 samples. This allowed us to estimate the probability *p*_VOC_ that a given SGTF sample was B.1.1.7 based upon its specimen date and NHS England region (**Extended Data Fig. 5**). Again restricting the analysis to specimens from 1 November 2020 onward, we find a hazard ratio associated with *p*_VOC_ of 1.58 (1.42–1.76) for the complete-cases analysis and 1.61 (1.42–1.82) for the IPW analysis (**Fig. 2c–d**).

## Absolute risks

To put these results into context, we calculated absolute mortality risks by applying hazard ratios for SGTF to the baseline risk of death among individuals tested in the community between August–October 2020 (assumed to be representative of the CFR associated with preexisting variants of SARS-CoV-2; **Table 1**). For the complete-cases analysis, in females aged 70–84, the estimated risk of death within 28 days of a positive SARS-CoV-2 test increases from 2.9% without SGTF to 4.4% with SGTF (95% CI 4.0–4.9%) and for females 85 or older increases from 13% to 19% (17–21%). For males aged 70–84 the risk of death within 28 days increases from 4.7% to 7.2% (6.4–7.9%) and for males 85 or older it increases from 17% to 25% (23– 27%). Estimates based on the IPW analysis corrected for misclassification were marginally higher. These estimates reflect a substantial increase in absolute risk amongst older age groups, but the risk of COVID-19 death following a positive test in the community remains below 1% in most individuals younger than 70 years old. Note that these estimates capture the fatality ratio among people tested in the community, and are thus likely to be higher than the infection fatality ratio, as many infected individuals are never tested.

**Table 1.** Absolute 28-day mortality risk associated with B.1.1.7, as expressed by case fatality ratio (%) among individuals testing positive in the community. The baseline risk (i.e., for preexisting SARS-CoV-2 variants) is derived using linked deaths within 28 days for all individuals testing positive in the community from 1 August – 31 October 2020. Adjusted risks are presented for the SGTF analysis for complete cases and for the misclassification-adjusted (*p*_VOC_) IPW analysis, which yielded the lowest and highest mortality estimates, respectively, of the main models assessed (**Fig. 2a–d**).

| Sex | Age | Baseline | SGTF, complete cases | $p_{voc}$, IPW |
| --- | --- | --- | --- | --- |
| Female | 0–34 | 0.00069% | 0.0011% (0.00096–0.0012%) | 0.0011% (0.00097–0.0012%) |
|  | 35–54 | 0.033% | 0.050% (0.045–0.056%) | 0.052% (0.046–0.059%) |
|  | 55–69 | 0.18% | 0.28% (0.25–0.31%) | 0.29% (0.26–0.33%) |
|  | 70–84 | 2.9% | 4.4% (4.0–4.9%) | 4.6% (4.0–5.1%) |
|  | 85 and older | 13% | 19% (17–21%) | 20% (18–22%) |
| Male | 0–34 | 0.0031% | 0.0047% (0.0042–0.0052%) | 0.0049% (0.0043–0.0055%) |
|  | 35–54 | 0.064% | 0.099% (0.089–0.11%) | 0.10% (0.090–0.12%) |
|  | 55–69 | 0.56% | 0.86% (0.77–0.95%) | 0.89% (0.78–1.0%) |
|  | 70–84 | 4.7% | 7.2% (6.4–7.9%) | 7.4% (6.6–8.3%) |
|  | 85 and older | 17% | 25% (23–27%) | 26% (23–29%) |

## Further investigations

We conducted a number of sensitivity analyses to verify the robustness of our results. Our main results were largely insensitive to: restriction to death-certificate-confirmed COVID-19 deaths only; any follow-up time of 21 days or longer; coarseness of geographical and temporal stratification; use of linear versus spline terms for age and IMD; analysis start date; follow-up time–covariate interactions; removal of the 10-day death registration cutoff; and restriction of the analysis to individuals with a full 28-day follow-up period (**Fig. 2e**). Generally, the IPW analysis yielded marginally higher hazard ratios, with greater uncertainty. As a further sensitivity analysis, we adjusted for an indicator in Pillar 2 testing data for whether the subject was tested because of symptoms or due to asymptomatic screening. Although we caution that symptomatic screening status may lie on the causal pathway between SGTF status and death, we found that this adjustment had no effect on the relative hazard of SGTF (1.54 [1.39–1.71], complete-cases analysis).

## Discussion

We previously found that B.1.1.7 is substantially more transmissible than preexisting SARS-CoV-2 variants, but could not robustly identify any associated change in disease severity using population-level analysis of early data^2^. This analysis of individual-level data, which controls for factors that could confound the association between B.1.1.7 infection and death, reveals an increase in COVID-19 mortality associated with lineage B.1.1.7. We stratify our analyses by test time and geographical location—mimicking matching on these variables—to account for changes in testing rates and changing pressures on hospital services over time and by region. Our findings are consistent with earlier reports^7^ by ourselves and other groups assessing the risk of death among individuals with SGTF. Crucially, our study is limited to individuals tested in the community. Indicators for B.1.1.7 infection are not currently available for most people who die from COVID-19 in England, as they are tested in the hospital rather than in the community and hospitals do not routinely collect genotypic data. However, this restricted focus allows us to capture the combined effect of an altered risk of hospitalisation given a positive test and an altered risk of death given hospitalisation, while only the latter would be measurable in a study of hospitalised patients only. Unfortunately, we were unable to account for vaccination status in this analysis.

We do not identify any mechanism for increased mortality here. B.1.1.7 infections are associated with higher viral concentrations on nasopharyngeal swabs, as measured by Ct values from PCR testing (**Extended Data Fig. 6**). Higher viral load could therefore be partly responsible for the observed increase in mortality; this could be assessed using a mediation analysis. Alternatively, changes in test-seeking behaviour could, in principle, explain our results. If B.1.1.7 infections were less likely to cause symptoms, but symptomatic B.1.1.7 cases were more severe, then our study could overestimate changes in the infection fatality rate. However, we find no clear difference in SGTF frequency among community tests relative to a random sample of SARS-CoV-2 infections in the population (**Extended Data Fig. 7**), suggesting that variant-associated changes in test-seeking propensity do not explain our findings.

## Data Availability

All analysis code is publicly available. An anonymised data set allowing replication of the analysis is publicly available.

https://github.com/nicholasdavies/cfrvoc

https://zenodo.org/record/4579857

## CMMID COVID-19 Working Group authors

The CMMID COVID-19 working group is (randomized order) Kevin van Zandvoort^1^, Samuel Clifford^1^, Fiona Yueqian Sun^1^, Sebastian Funk^1^, Graham Medley^1^, Yalda Jafari^1^, Sophie R Meakin^1^, Rachel Lowe^1^, W John Edmunds^1^, Matthew Quaife^1^, Naomi R Waterlow^1^, Rosalind M Eggo^1^, Nicholas G. Davies^1^, Jiayao Lei^1^, Mihaly Koltai^1^, Fabienne Krauer^1^, Damien C Tully^1^, James D Munday^1^, Alicia Showering^1^, Anna M Foss^1^, Kiesha Prem^1^, Stefan Flasche^1^, Adam J Kucharski^1^, Sam Abbott^1^, Billy J Quilty^1^, Thibaut Jombart^1^, Alicia Rosello^1^, Gwenan M Knight^1^, Mark Jit^1^, Yang Liu^1^, Jack Williams^1^, Joel Hellewell^1^, Kathleen O’Reilly^1^, Yung-Wai Desmond Chan^1^, Timothy W Russell^1^, Christopher I Jarvis^1^, Simon R Procter^1^, Akira Endo^1^, Emily S Nightingale^1^, Nikos I Bosse^1^, C Julian Villabona-Arenas^1^, Frank G Sandmann^1^, Amy Gimma^1^, Kaja Abbas^1^, William Waites^1^, Katherine E. Atkins^1,4^, Rosanna C Barnard^1^, Petra Klepac^1^, Hamish P Gibbs^1^, Carl A B Pearson^1^, and Oliver Brady^1^.

4. Centre for Global Health, Usher Institute of Population Health Sciences and Informatics, University of Edinburgh, Edinburgh, UK.

## Methods

### Ethical approval

Approved by the Observational / Interventions Research Ethics Committee at the London School of Hygiene and Tropical Medicine (reference number 24020). Subject consent is not required for national infectious disease notification data sets in England.

### Data sources

We linked three datasets provided by Public Health England: a line list of all positive tests in England’s “Pillar 2” (community) testing for SARS-CoV-2, containing specimen date and demographic information on the test subject; a line list of cycle threshold (Ct) values for the ORF1ab, N (nucleocapsid), and S (spike) genes for positive tests that were processed in one of the three national laboratories (Alderley Park, Glasgow, or Milton Keynes) utilising the Thermo Fisher TaqPath COVID-19 assay; and a line list of all deaths due to COVID-19 in England, which combines and deduplicates deaths reported by hospitals in England, by the Office for National Statistics, via direct reporting from Public Health England Health Protection Team, and via Demographic Batch Service tracing of laboratory-confirmed cases^8^. We link these datasets using a numeric identifier for Pillar 2 tests (‘FINALID’) common to all three datasets. We define S gene target failure (SGTF) as any test with Ct < 30 for ORF1ab and N targets but no detectable S gene, and non-SGTF as any test with Ct < 30 for ORF1ab, N, and S targets. A small proportion (10.4%) of SGTF tests are inconclusive. The study population of interest is defined as all individuals who received a positive Pillar 2 test between 1 November 2020 and 14 February 2021. For our main analysis, we included only tests from after 1 November 2020 to avoid including an excess of tests with SGTF not resulting from infection by lineage B.1.1.7. In sensitivity analyses, we also consider extending the population to include tests performed between 1 September and 31 October 2020.

Our analysis does not include individuals who first tested positive in hospital, that is, those who presented to hospital after symptom onset without first being tested in the community. This is because cycle threshold values used to ascertain SGTF status are not available for individuals who were not tested in the community. Of the 57,750 COVID-19 deaths in England during the study period, 17,642 deaths (44%) can be linked to a positive Pillar 2 test; among these, 4,945 have non-missing SGTF status. So, while our study includes 1,098,729 Pillar 2 tests with non-missing SGTF status, which represents 51.1% of the 2,245,263 Pillar 2 tests over this period and 40.2% of the 2,736,806 combined Pillar 1 (hospital) and Pillar 2 (community) SARS-CoV-2 tests over this period, we can only assess SGTF status for 9% (4,945 / 57,750) of the individuals who died from COVID-19 over the study period. This is explained by differing mortality rates among individuals who first test positive in a hospital compared to those who are tested in the community, as the former group are much more likely to have severe illness, as well as by missingness in the SGTF data.

There was a small amount of missing data for sex (*n* = 14, <0.01%), age (*n* = 171, <0.01%), and IMD and regional covariates (*n* = 3,817, 0.16%). There were no missing specimen dates. Individuals with missing age, sex, or geographical location were excluded. We also excluded individuals from the dataset whose age was recorded as zero, as there were 17,913 age-0 individuals compared to 10,132 age-1 individuals in the dataset, suggesting that many of these age-0 individuals may have been miscoded. There was some missing data on ethnicity (*n* = 47,491, 2%) and we created a category that combines missing values with “Other” and “Mixed”. Missing values for residence type (*n* = 63,905, 3%) were also combined with an “Other” category. The full data set used for the main analysis comprises 2,245,263 individuals, with SGTF status missing or inconclusive for 1,098,729 (48.9%). Missing data on the exposure is addressed in the analysis, described below.

We grouped residence types into three categories: Residential, which included the “Residential dwelling (including houses, flats, sheltered accommodation)” and “House in multiple occupancy (HMO)” groups; Care/Nursing home; and Other/Unknown, which included the “Medical facilities (including hospitals and hospices, and mental health)”, “No fixed abode”, “Other property classifications”, “Overseas address”, “Prisons, detention centres, secure units”, “Residential institution (including residential education)”, and “Undetermined” groups, as well as unspecified residence type. We grouped ethnicities into four categories according to the broad categories used in the 2011 UK Census: Asian, which included the “Bangladeshi (Asian or Asian British)”, “Chinese (other ethnic group)”, “Indian (Asian or Asian British)”, “Pakistani (Asian or Asian British)”, and “Any other Asian background” groups; Black, which included the “African (Black or Black British)”, “Caribbean (Black or Black British)”, and “Any other Black background” groups; White, which included the “British (White)”, “Irish (White)”, and “Any other White background” groups; and Other / Mixed / Unknown, which included the “Any other ethnic group”, “White and Asian (Mixed)”, “White and Black African (Mixed)”, “White and Black Caribbean (Mixed)”, “Any other Mixed background”, and “Unknown” groups.

### Statistical methods

There are several factors that we expect to be associated with both SGTF and with risk of death, thus confounding the association between SGTF and risk of death in those tested. Area of residence and specimen date were expected to be potentially strong confounders. Area of residence is expected to be strongly associated with SGTF status due to different virus variants circulating in different areas, and specimen date because the prevalence of SGTF is known to have greatly increased over time. Area of residence and specimen date are also expected to be associated with risk of death following a test, including due to differential pressure on hospital resources by area and time. The following variables were also identified as potential confounders: sex, age, place of residence (Residential, Care/Nursing home, or Other/Unknown), ethnicity (White, Asian, Black, or Other/Mixed/Unknown), index of multiple deprivation (IMD). The potential confounders are referred to collectively as the covariates. For descriptive analyses, age (in years) was categorised as 1–34, 35–54, 55–69, 70–84, or 85 and older.

Descriptive analyses were performed. We tabulated the distribution of the covariates in the whole study sample, the association between each covariate and SGTF status in the subset with SGTF measured, and the association between each covariate and missing data in SGTF status **(Extended Data Table 1)**. The subset with SGTF status measured are referred to as the complete cases. The unadjusted association between SGTF and mortality in the complete cases was assessed using a Kaplan-Meier plot (**Fig. 1c**), and Kaplan-Meier plots and crude 28-day mortality rates are also presented separately according to categories of the covariates (**Extended Data Table 2, Extended Data Fig. 2**). Crude overall mortality rates (i.e., not restricted to 28 days after a positive test) were obtained for the whole sample, by SGTF status in the complete cases, and in those with missing SGTF status, according to categories of each covariate (**Supplementary Table 1**). We also obtained mortality rates by SGTF status (in the complete cases) for categories of each covariate stratified by age group (**Fig. 1d–i**). Exact Poisson CIs are used for mortality rates, assuming constant rates.

Approximately 49% of individuals in the study sample are missing data on SGTF status, due to their test not being sent to one of the three laboratories utilising the Thermo Fisher TaqPath COVID-19 assay or the test being inconclusive. We performed complete cases analysis, restricted to the subset with SGTF status measured. This complete case analysis assumes that for each analysis, the missing data, in this case missing SGTF status, is independent from the outcome of interest, given the variables included in the models. This is a specific type of missing not at random (MNAR) assumption, as in particular it is allowed to depend on the underlying value of SGTF. We also performed an analysis of the complete cases using inverse probability weights^5^ (IPW) to address the missing data on SGTF, under a missing at random (MAR) assumption. In the analysis, each individual with SGTF status measured is weighted by the inverse of their probability of having SGTF status measured based on their covariates. For the IPW, the missingness model estimated the probability of missingness using logistic regression with age (restricted cubic spline), sex, IMD decile (restricted cubic spline), ethnicity, residence type by asymptomatic screening indicator, and NHS region by specimen week as predictors. We also considered a cauchit and a Gosset link for the missingness model, including the same predictors, as this was expected to provide better stability for the weights^5^. The fit of the missingness model was assessed using a Q-Q plot (**Extended Data Fig. 4**), and Hosmer-Lemeshow and Hinkley tests were used to choose the most appropriate model.

Cox regression^4^ was used to estimate the association between SGTF and the hazard of mortality, conditioning on the potential confounders listed above. The analyses described here were applied to the complete cases and using IPW. For IPW analyses, the standard errors (SEs) accounted for the weights, though the fact that the weights were estimated was not accounted for; this results in conservative SEs. The baseline hazard in the Cox model was stratified by both specimen date and LTLA, therefore finely controlling for these variables. The stratification gives a large number of strata matched by specimen date and LTLA. Only those strata that contain individuals who die and individuals who survive contribute to the analysis. The analysis is therefore similar to that which would be performed had we created a matched nested case-control sample. The remaining variables were included as covariates in the model (sex, age, place of residence, ethnicity, IMD decile). Age was included as a restricted cubic spline with 5 knots, and IMD decile was included as a restricted cubic spline with 3 knots. The time origin for the analysis was specimen date and we considered deaths up to 28 days after the specimen date. Individuals who did not die within 28 days were censored at the earlier of 28 days post specimen date and the administrative censoring date, which we chose as the date of the most recent death linkable to SGTF status minus 10 days (i.e., 14 February 2021) in order to minimise any potential bias due to late reporting of deaths. We began by assuming proportionality of hazards for SGTF and the covariates included in the model. The proportional hazards assumption was assessed by including in the model an interaction between each covariate and time, which was performed separately for SGTF and for each other covariate. Schoenfeld residual plots were also obtained for each covariate (**Extended Data Fig. 3**). We assessed whether the association between SGTF and the hazard was modified by age, sex, IMD, ethnicity, and place of residence. Models with and without interactions were compared using likelihood ratio tests for the complete cases analyses. For the analysis using IPW we used Wald tests based on robust standard errors^9^.

The analysis assumes that censoring is uninformative, which is plausible as all censoring is administrative.

### Misclassification analysis

The exposure of SGTF is subject to misclassification, because a number of minor circulating SARS-CoV-2 lineages in addition to B.1.1.7 are also associated with failure to amplify the spike gene target. Accordingly, a positive test with SGTF is not necessarily indicative of infection with B.1.1.7. A negative test of SGTF is assumed to be indicative of absence of infection with B.1.1.7. Misclassification of an exposure can result in bias in its estimated association with the outcome. We fitted a logistic model to Pillar 2 SGTF frequencies by NHS region to estimate a “background” rate of SGTF in the absence of B.1.1.7, assuming a beta binomial prior. This model is then used to estimate the probability that an individual testing positive with SGTF is infected with B.1.1.7, separately for individuals in each NHS region. These probabilities can then be used in place of the indicator of SGTF exposure in the Cox models. This is the regression calibration approach^6^ to correcting for bias due to measurement error in an exposure.

We fitted models accounting for false positives (modelled as regionally-varying background rates of SGTF associated with non-B.1.1.7 variants) to the SGTF data. Our logistic model for B.1.1.7 growth over time is as follows:

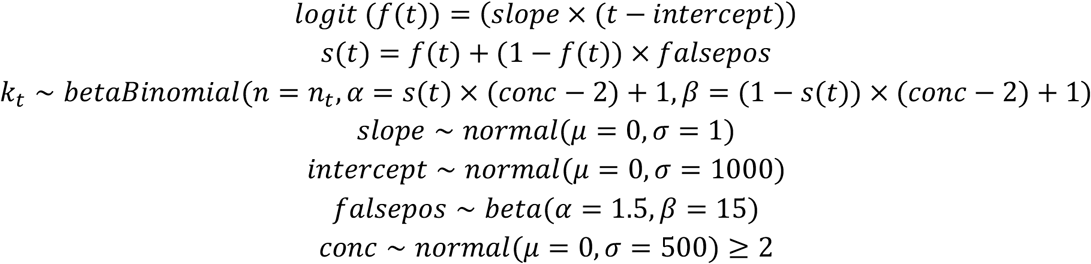

Here, *f*(*t*) is the predicted frequency of B.1.1.7 among positive tests at time *t* (in days since 1 September 2020) based on the terms *slope* and *intercept*; *s*(*t*) is the predicted frequency of S gene target failure at time *t* due to the combination of B.1.1.7 and a background false positive rate *falsepos, conc* is the “concentration” parameter (= α + β) of a beta distribution with mode *s*(*t*); *k*_*t*_ is the number of S gene target failures detected at time *t*; and *n*_*t*_ is the total number of tests at time *t*. All priors above are chosen to be vague, and the truncation of *conc* to values greater than 2 ensures a unimodal distribution for the proportion of tests that are SGTF. The model above is fitted separately for each NHS England region. Then, *p*_VOC_ for a test with SGTF = 1 at time *t* is equal to *f(t)/s(t)*, and *p*_VOC_ = 0 for all tests with SGTF = 0. The model was fitted using Markov chain Monte Carlo with 10,000 iterations of burn-in and 5,000 iterations of sampling.

The model above was fitted using the same data source (i.e. SGTF frequencies among Pillar 2 community tests for SARS-CoV-2) as our survival analysis. To verify the robustness of this model, we performed a sensitivity analysis using sequencing data from the COVID-19 UK Genomics Consortium^10^ downloaded from the Microreact platform^11^ on 11 January 2020 to estimate *p*_VOC_. In this alternative analysis we estimated *p*_VOC_ for each NHS England region and date as the number of samples that were VOC 202012/01 (i.e. lineage B.1.1.7 with mutations Δ69/Δ70 and N501Y in Spike) divided by the number of samples that were SGTF (i.e. any lineage with Δ69/Δ70, the deletion that causes SGTF) for that NHS England region and date, setting *p*_VOC_ = 1 for all dates later than 31 December 2020 as there were no sequencing data available past this date, and filling any gaps in the data using linear interpolation. This yielded nearly identical results in our survival analysis compared to using the modelled *p*_VOC_ described above (**Fig. 2e**).

### Absolute risks

Estimates from the final Cox models were used to obtain estimates of absolute risk of death for 28 and 60 days with SGTF and *p*_VOC_. Given the strong influence of age on risk of death, we present absolute risks by sex and age group (1–34, 35–54, 55–69, 70–84, 85+). Absolute risks of death (case fatality rate) within 28 days were estimated by age group and sex using data on individuals tested during August–October 2020; this is referred to as the baseline risk. The absolute risks of death for individuals with SGTF were then estimated as follows. If the baseline absolute risk of death in a given age group is 1− *A*, then the estimated absolute risk of death with SGTF is 1−*A*^*HR*^, where HR denotes the estimated hazard ratio obtained from the Cox model assuming proportional hazards. Standard errors are obtained via the delta method, and CIs based on normal approximations.

### Sensitivity analyses

Several sensitivity analyses were performed. After establishing the final model through using the process outlined above, we investigated the impact of using different variables for stratification of the baseline hazard measuring region at a coarser level (UTLA, or NHS England region), as well as coarser test specimen time (week rather than exact date). Adjusting for these variables instead of using stratification was also explored. We also repeated the main analysis restricting data to specimens collected from September onwards, October onwards, November onwards, or December onwards.

To assess the impact of imposing an administrative cutoff to follow-up time of 10 days prior to data extraction, we first reanalysed the data without this cutoff, as well as reanalysing the data restricting the analysis to individuals with at least 28 days’ follow-up.

Finally, we adjusted for symptomatic status associated with the test (asymptomatic versus symptomatic), which relates to whether the test was given for asymptomatic screening purposes or on the basis of a request by a (presumed symptomatic) individual, as only symptomatic individuals may request a community SARS-CoV-2 test in England.

## Acknowledgements

We gratefully acknowledge the assistance of Public Health England in providing the analysis data and authorising release of an anonymised data set. This work was supported by grants from UK Research and Innovation, the National Institutes of Health Research (NIHR) Health Protection Research Unit in Immunisation [NIHR200929], and the UK Medical Research Council [MC_PC_19065] (NGD); Global Challenges Research Fund project ‘RECAP’ managed through Research Councils UK and the Economic and Social Research Council [ES/P010873/1] (CIJ); European Commission project ‘EpiPose’ [101003688] and the NIHR [NIHR200908] (WJE); the National Institutes of Health / National Institute of Allergy and Infectious Diseases [R01AI148127] (NPJ); a Royal Society-Wellcome Trust Sir Henry Dale Fellowship [218554/Z/19/Z] (KDO); and a UKRI Future Leaders Fellowship [MR/S017968/1] (RHK). See Supplementary Note 2 for working group acknowledgements.

## Author contributions

NGD, CIJ, KDO and RHK performed analyses; all authors designed the study and wrote the manuscript.

## Additional information

Correspondence should be addressed to Nicholas G. Davies.

## Data availability

An anonymised data set allowing replication of the analysis is available at https://zenodo.org/record/4579857.

## Code availability

Analysis code deposited at time of publication is available at https://zenodo.org/record/4579857. The repository is maintained at https://github.com/nicholasdavies/cfrvoc.

## Supplementary Information

Supplementary Tables 1–2; Supplementary Notes 1–2.

## Extended Data

**Extended Data Table 1.**
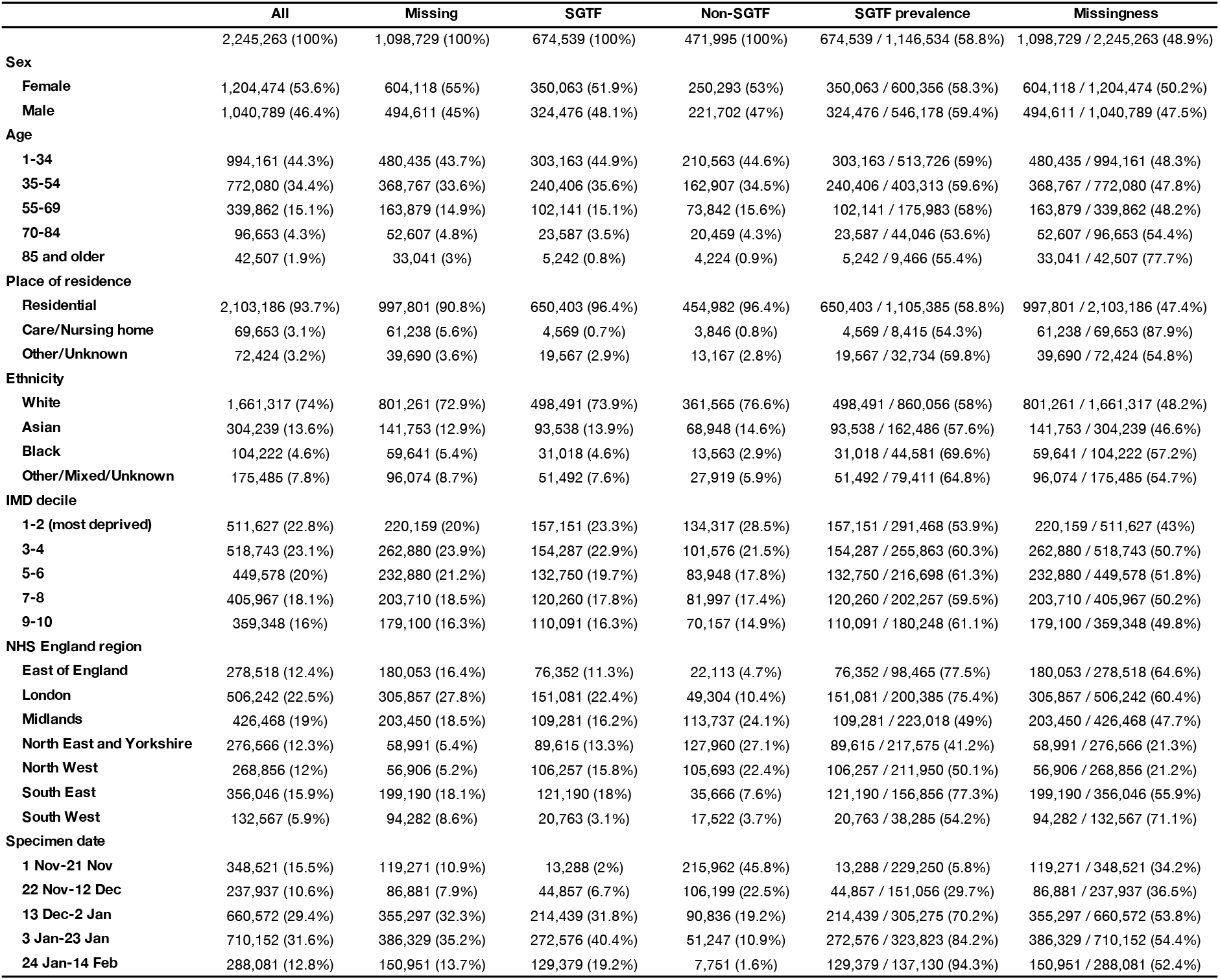
Characteristics of study subjects, 1 November 2020–14 February 2021. **All**, N (%); **Missing**, N (%); **SGTF**, N with SGTF (%) in subset of tests with non-missing SGTF status; **Non-SGTF**, N with non-SGTF (%) in subset of tests with non-missing SGTF status; **SGTF prevalence**, N with SGTF / total (%) in subset with known SGTF status; **Missingness**, N with missing SGTF status / total (%).

**Extended Data Table 2.**
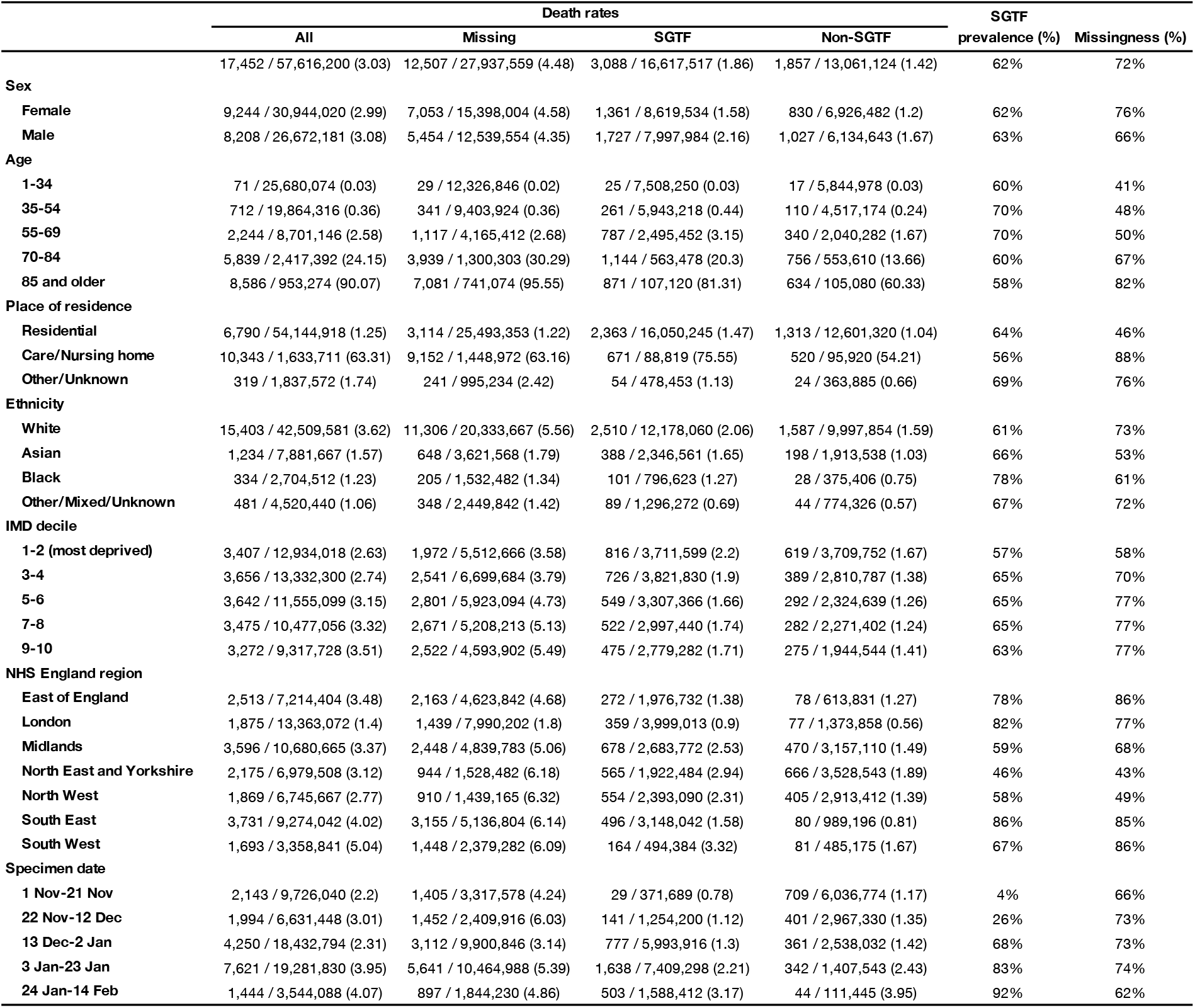
Rates of death within 28 days of positive test among study subjects. Total number of deaths, number of days of followup, and deaths per 10,000 days of followup reported for: **All** deaths, **Missing** SGTF status deaths, known **SGTF** deaths and known **Non-SGTF** deaths. Missingness among deaths (%) and SGTF prevalence among deaths with non-missing SGTF status (%) are also reported.

**Extended Data Fig. 1.**
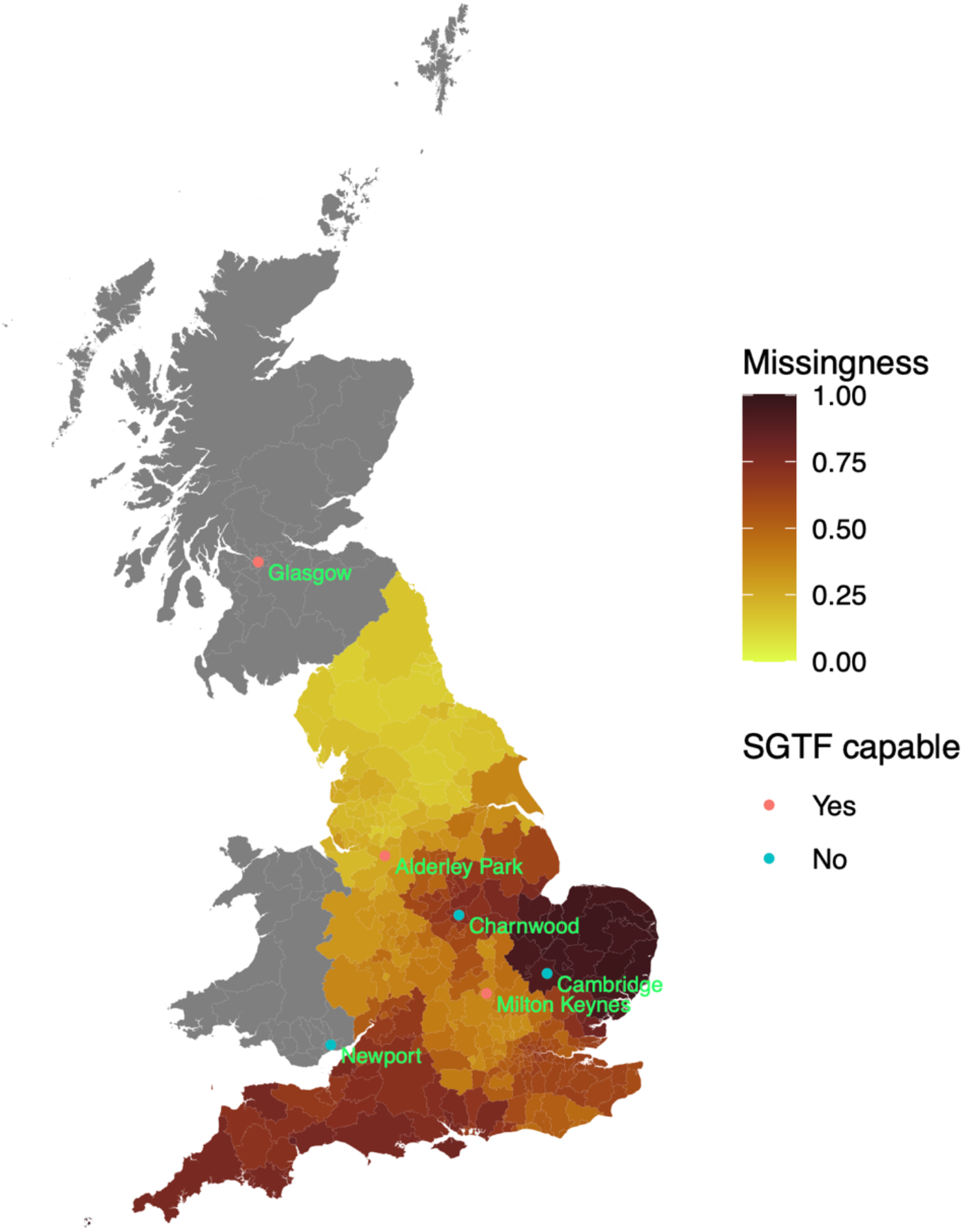
Missingness in SGTF status and proximity to SGTF-capable Lighthouse lab. The geographical location of the six Lighthouse Labs in the United Kingdom; missingness is higher in the lower-tier local authorities (shaded regions) which are closer to a Lighthouse lab that is not capable of producing an SGTF reading.

**Extended Data Fig. 2.**
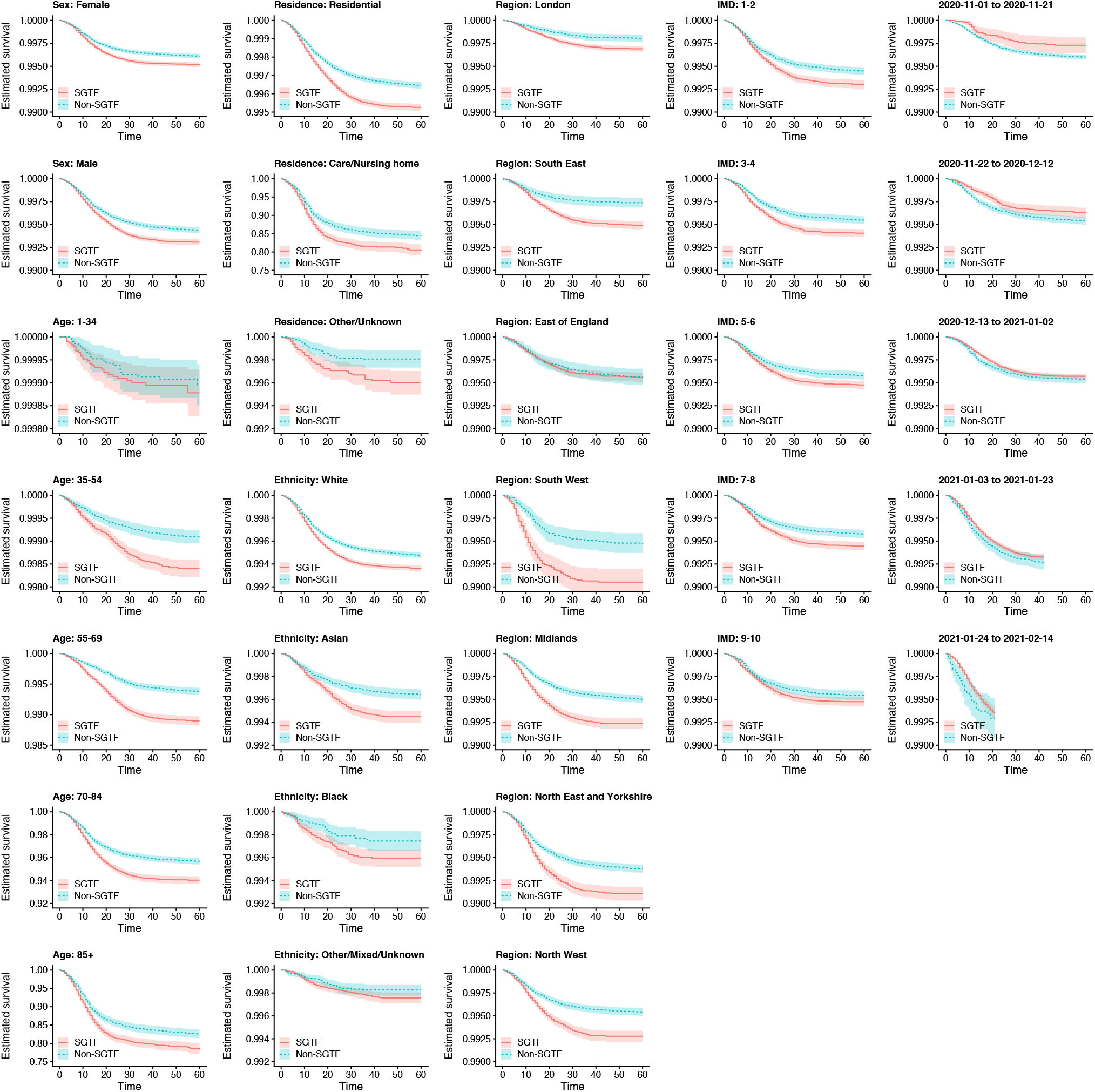
Kaplan-Meier plots of survival within 60 days of positive test for SGTF versus non-SGTF. Plots are stratified by sex, age group, place of residence, ethnicity, NHS England region, IMD decile (in 5 groups), and specimen date. Note that *y*–axis ranges differ among panels. These curves show the crude survival within each group (unadjusted for other covariates), and so do not necessarily signify differences in the effect of SGTF on survival for any specific group due to possible confounding. Shaded areas show 95% CIs.

**Extended Data Fig. 3.**
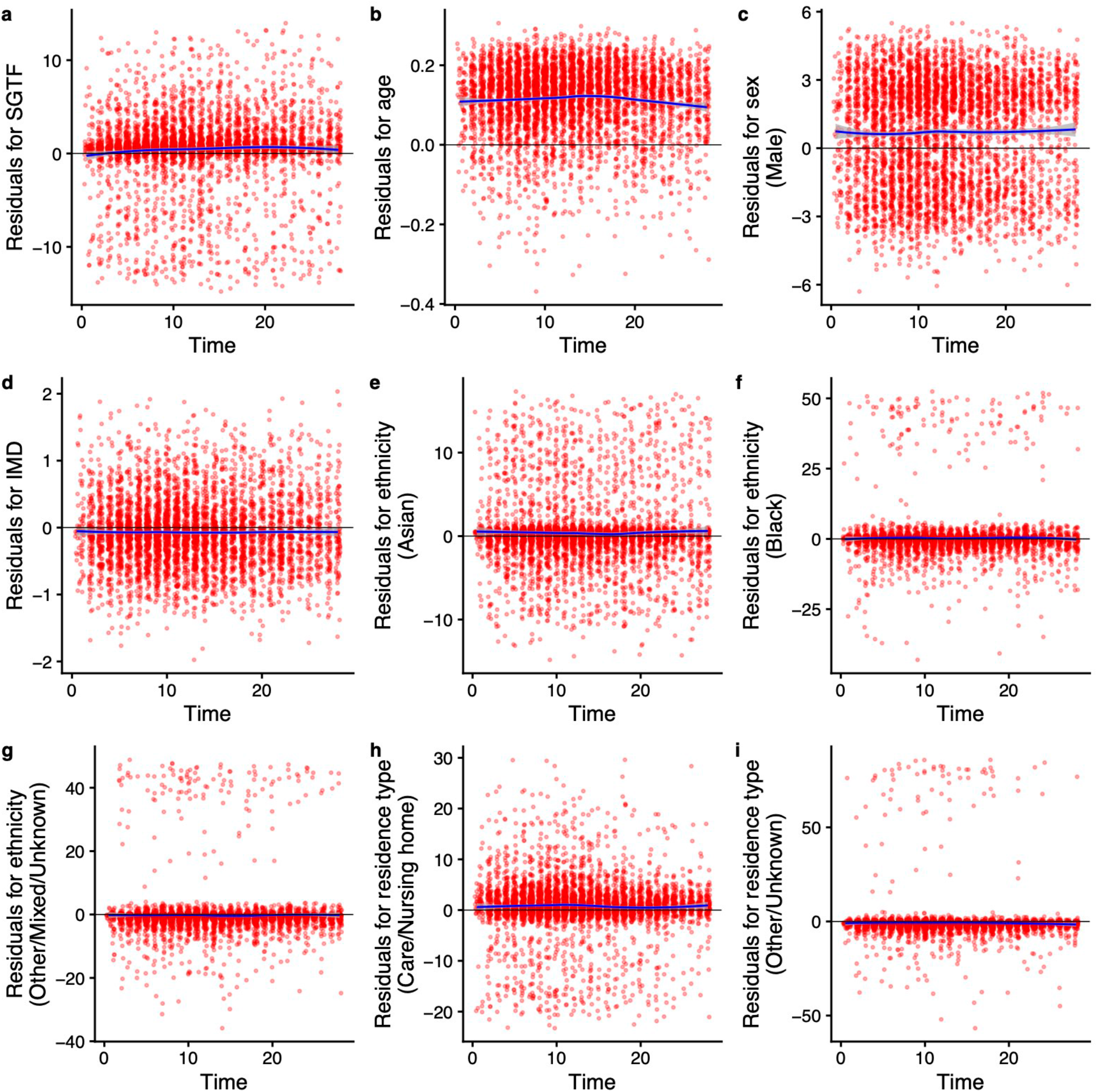
Schoenfeld residuals for survival model by SGTF stratified by LTLA and specimen date. Model uses linear terms for age and IMD and a 28-day followup using complete cases. Schoenfeld residual tests (two-sided) give (**a**) *P* = 0.001 for SGTF; (**b**) *P* = 0.039 for age; (**c**) *P* = 0.101 for sex; (**d**) *P* = 0.937 for IMD decile; (**e**–**g**) *P* = 0.969 for ethnicity; (**h**–**i**) *P* = 0.064 for residence type; and *P* = 0.027 globally. Trend line shows mean and 95% CIs of a loess regression.

**Extended Data Fig. 4.**
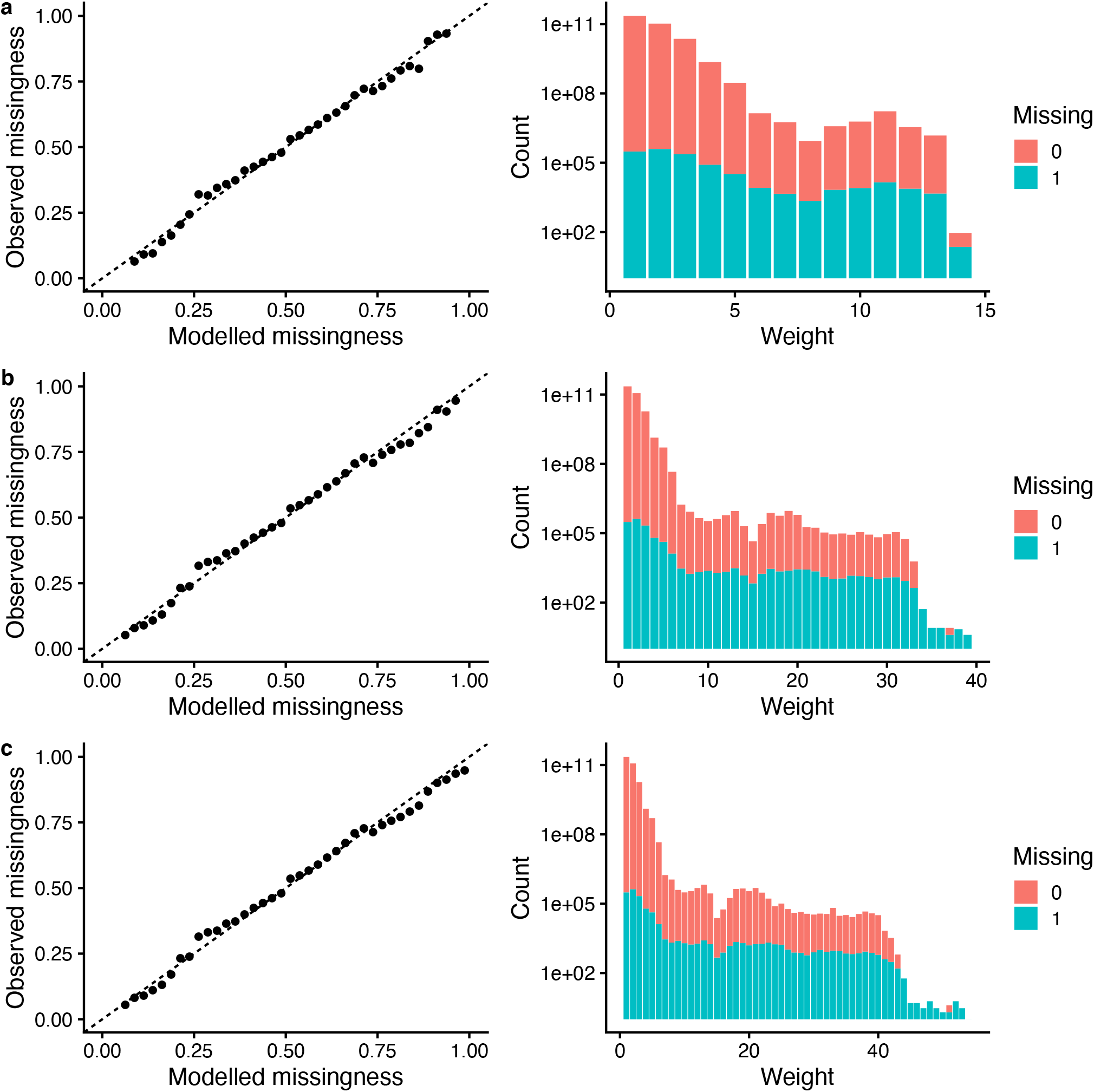
Comparison of missingness models. Q-Q plot (left; mean and 95% CIs) and distribution of weights (right) under different missingness models assessed for IPW with **(a)** a cauchit link, **(b)** a robit link (Student’s *t* distribution with d.f. = 4), and **(c)** a logit link.

**Extended Data Fig. 5.**
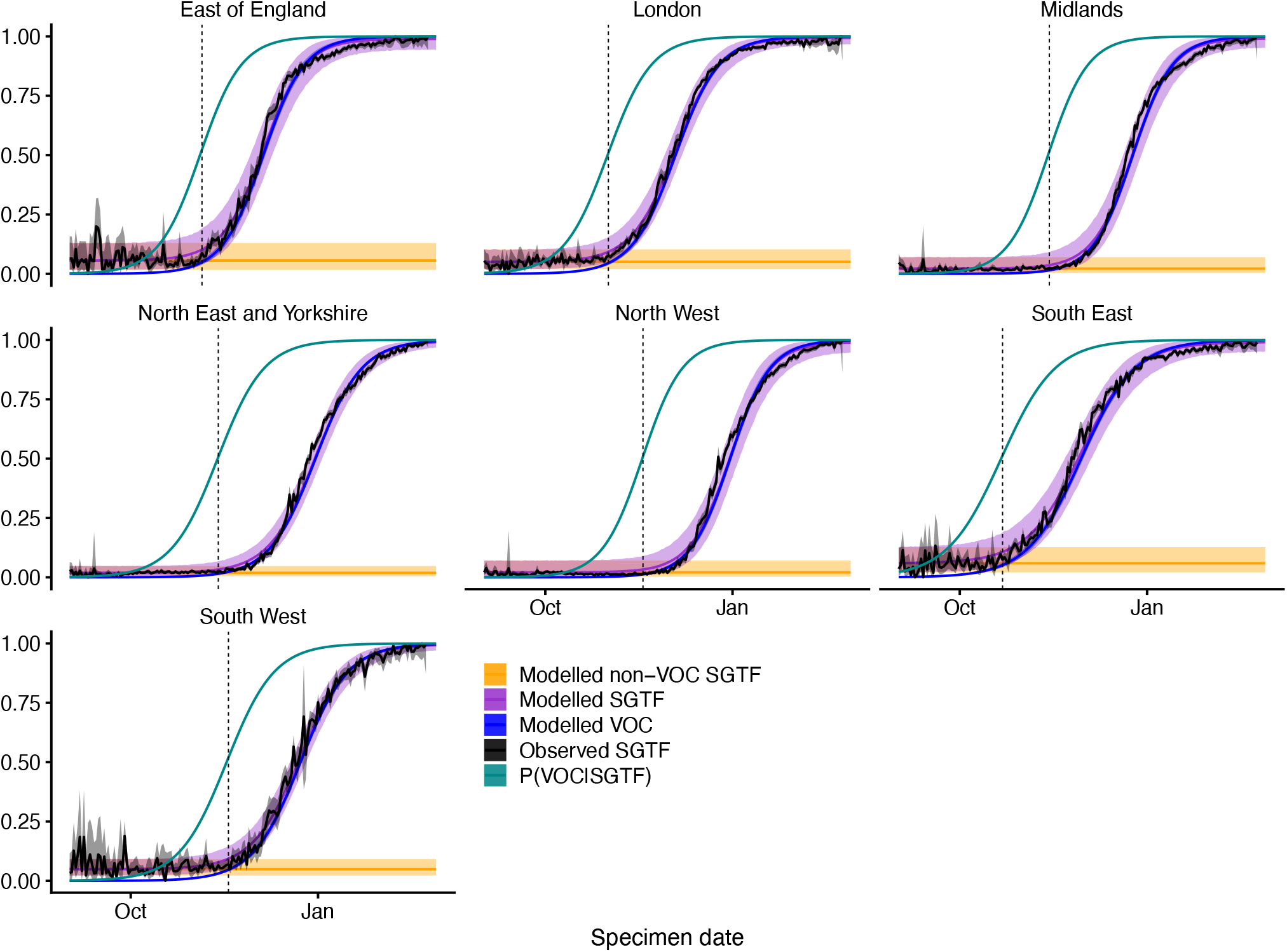
Misclassification model. For each NHS England region, we fit a beta-binomial model (purple, Modelled SGTF) to the observed SGTF frequencies among Pillar 2 tests (black, Observed SGTF), which estimates a constant proportion of “false positive” SGTF samples among non-VOC 202012/01 (i.e., non-B.1.1.7) specimens (orange, Modelled non-VOC SGTF) and a logistically growing proportion of VOC 202012/01 (i.e., B.1.1.7) specimens over time (blue, Modelled VOC). This allows us to model the conditional probability that a specimen with SGTF represents VOC 202012/01 (teal, P(VOC|SGTF)). For our misclassification survival analysis, *p*_VOC_ = 0 for non-SGTF specimens and *p*_VOC_ = P(VOC|SGTF) for SGTF specimens. Lines show medians and shaded areas show 95% credible intervals.

**Extended Data Fig. 6.**
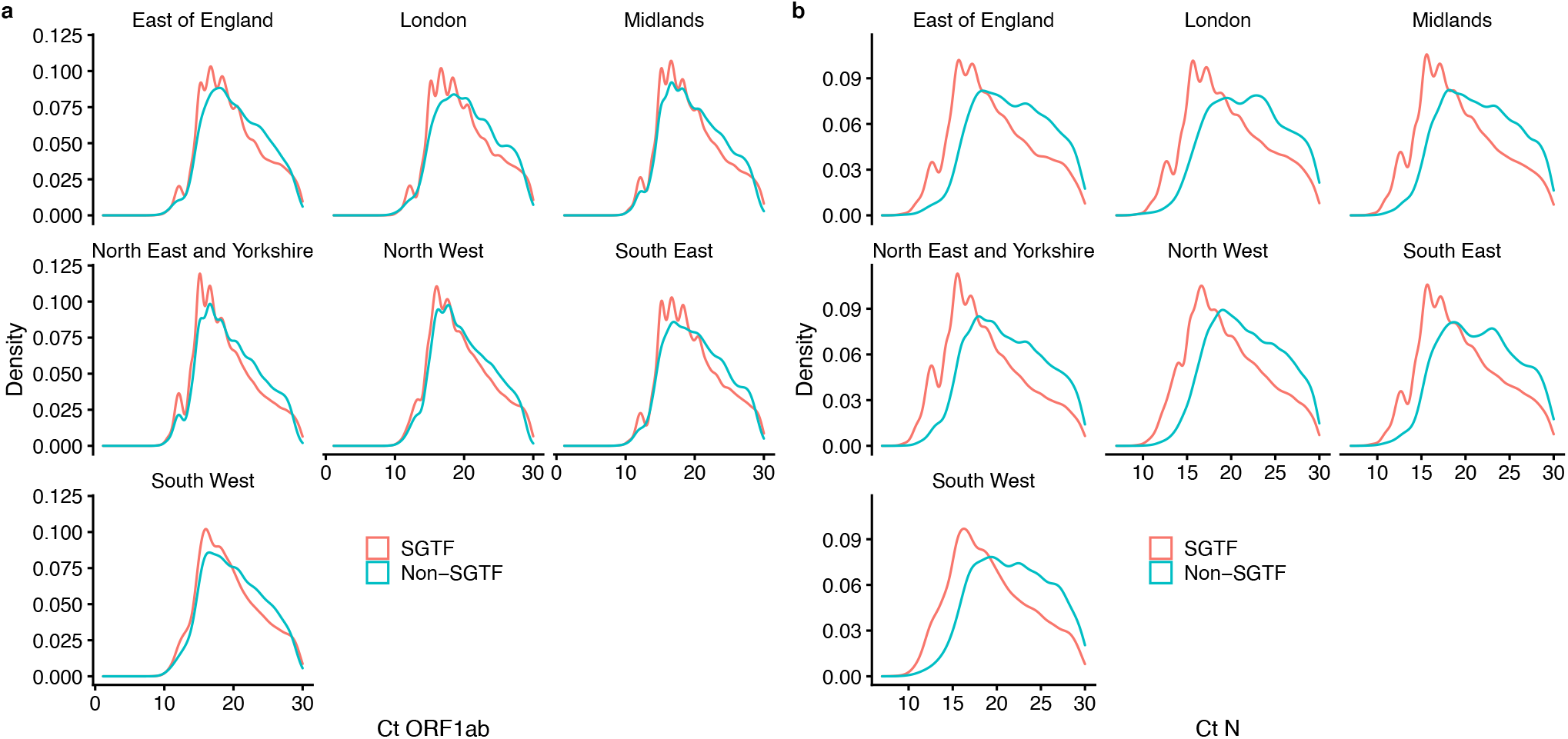
Ct values for SGTF versus non-SGTF. The distribution of Ct values for (**a**) ORF1ab and (**b**) N gene targets among specimens collected between 1 January–14 February 2021.

**Extended Data Fig. 7.**
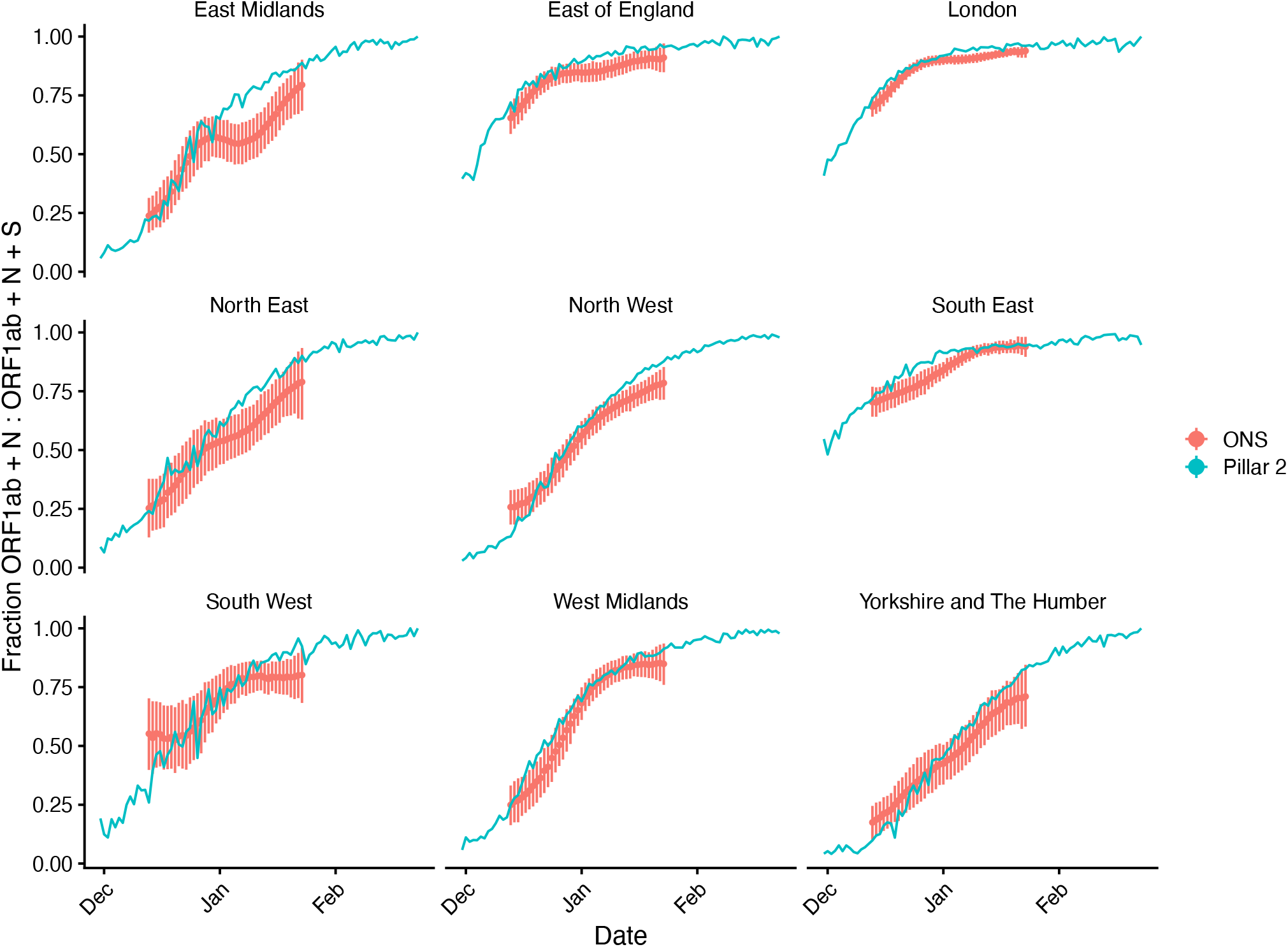
S gene dropout in community tests relative to a random sample of SARS-CoV-2 infections in the community. Comparison of the proportion of samples with S gene dropout in the Pillar 2 (i.e., community testing) sample used in this analysis compared to Office for National Statistics (ONS) random sampling of the community. This comparison suggests that S gene dropout samples are not overrepresented in testing data relative to the prevalence of S gene dropout in the community, suggesting that the increased hazard of death among positive community tests estimated in this study is not the result of a decrease in the average propensity for test-seeking among individuals infected with B.1.1.7. Point and ranges for ONS data show mean and 95% credible intervals.

## Supplementary Information for

**Supplementary Table 1.** Rates of death within any time period following positive test among study subjects, including missing SGTF status. Total number of deaths, number of days of followup, and deaths per 10,000 days of followup reported. The maximum observed followup was 105 days.

|  | All | Missing | SGTF | Non-SGTF |
| --- | --- | --- | --- | --- |
|  | 19,615 / 111,457,728 (1.76) | 13,989 / 50,338,187 (2.78) | 3,362 / 25,927,734 (1.3) | 2,264 / 35,191,806 (0.64) |
| **Sex** |  |  |  |  |
| Female | 10,393 / 59,964,454 (1.73) | 7,923 / 27,867,530 (2.84) | 1,461 / 13,457,980 (1.09) | 1,009 / 18,638,942 (0.54) |
| Male | 9,222 / 51,493,274 (1.79) | 6,066 / 22,470,656 (2.7) | 1,901 / 12,469,754 (1.52) | 1,255 / 16,552,864 (0.76) |
| **Age** |  |  |  |  |
| 1-34 | 84 / 50,157,768 (0.02) | 32 / 22,443,774 (0.01) | 29 / 11,879,958 (0.02) | 23 / 15,834,034 (0.01) |
| 35-54 | 856 / 38,321,232 (0.22) | 398 / 16,839,060 (0.24) | 309 / 9,307,178 (0.33) | 149 / 12,174,994 (0.12) |
| 55-69 | 2,641 / 16,670,620 (1.58) | 1,285 / 7,431,238 (1.73) | 903 / 3,767,472 (2.4) | 453 / 5,471,911 (0.83) |
| 70-84 | 6,528 / 4,629,068 (14.1) | 4,413 / 2,335,022 (18.9) | 1,215 / 832,234 (14.6) | 900 / 1,461,813 (6.16) |
| 85 and older | 9,506 / 1,679,038 (56.62) | 7,861 / 1,289,092 (60.98) | 906 / 140,892 (64.3) | 739 / 249,054 (29.67) |
| **Place of residence** |  |  |  |  |
| Residential | 7,796 / 104,825,376 (0.74) | 3,550 / 45,778,629 (0.78) | 2,606 / 25,079,712 (1.04) | 1,640 / 33,967,036 (0.48) |
| Care/Nursing home | 11,472 / 3,134,891 (36.59) | 10,183 / 2,792,355 (36.47) | 692 / 115,209 (60.06) | 597 / 227,327 (26.26) |
| Other/Unknown | 347 / 3,497,460 (0.99) | 256 / 1,767,203 (1.45) | 64 / 732,813 (0.87) | 27 / 997,444 (0.27) |
| **Ethnicity** |  |  |  |  |
| White | 17,272 / 82,953,286 (2.08) | 12,635 / 37,068,638 (3.41) | 2,704 / 18,966,734 (1.43) | 1,933 / 26,917,914 (0.72) |
| Asian | 1,417 / 15,309,440 (0.93) | 731 / 6,447,530 (1.13) | 439 / 3,648,430 (1.2) | 247 / 5,213,480 (0.47) |
| Black | 377 / 4,801,714 (0.79) | 229 / 2,569,738 (0.89) | 113 / 1,251,373 (0.9) | 35 / 980,604 (0.36) |
| Other/Mixed/Unknown | 549 / 8,393,288 (0.65) | 394 / 4,252,282 (0.93) | 106 / 2,061,196 (0.51) | 49 / 2,079,810 (0.24) |
| **IMD decile** |  |  |  |  |
| 1-2 (most deprived) | 3,871 / 25,401,898 (1.52) | 2,239 / 10,049,132 (2.23) | 884 / 5,407,510 (1.63) | 748 / 9,945,257 (0.75) |
| 3-4 | 4,095 / 25,376,552 (1.61) | 2,831 / 11,867,324 (2.39) | 792 / 5,947,716 (1.33) | 472 / 7,561,513 (0.62) |
| 5-6 | 4,091 / 22,075,895 (1.85) | 3,128 / 10,566,596 (2.96) | 599 / 5,256,524 (1.14) | 364 / 6,252,776 (0.58) |
| 7-8 | 3,892 / 20,384,284 (1.91) | 2,966 / 9,458,600 (3.14) | 573 / 4,782,278 (1.2) | 353 / 6,143,406 (0.57) |
| 9-10 | 3,666 / 18,219,098 (2.01) | 2,825 / 8,396,536 (3.36) | 514 / 4,533,706 (1.13) | 327 / 5,288,855 (0.62) |
| **NHS England region** |  |  |  |  |
| East of England | 2,748 / 12,982,012 (2.12) | 2,347 / 8,084,884 (2.9) | 300 / 3,274,839 (0.92) | 101 / 1,622,290 (0.62) |
| London | 2,122 / 24,247,196 (0.88) | 1,602 / 13,541,876 (1.18) | 423 / 6,941,281 (0.61) | 97 / 3,764,038 (0.26) |
| Midlands | 4,091 / 21,368,735 (1.91) | 2,790 / 9,120,507 (3.06) | 723 / 3,789,278 (1.91) | 578 / 8,458,950 (0.68) |
| North East & Yorkshire | 2,536 / 15,708,002 (1.61) | 1,133 / 3,225,434 (3.51) | 599 / 2,654,326 (2.26) | 804 / 9,828,241 (0.82) |
| North West | 2,151 / 13,480,134 (1.6) | 1,066 / 2,839,457 (3.75) | 594 / 3,087,830 (1.92) | 491 / 7,552,846 (0.65) |
| South East | 4,125 / 17,202,003 (2.4) | 3,474 / 9,072,312 (3.83) | 552 / 5,446,734 (1.01) | 99 / 2,682,958 (0.37) |
| South West | 1,842 / 6,469,646 (2.85) | 1,577 / 4,453,717 (3.54) | 171 / 733,445 (2.33) | 94 / 1,282,484 (0.73) |
| **Specimen date** |  |  |  |  |
| 1 Nov-21 Nov | 2,830 / 33,119,124 (0.85) | 1,862 / 11,270,923 (1.65) | 37 / 1,243,146 (0.3) | 931 / 20,605,054 (0.45) |
| 22 Nov-12 Dec | 2,481 / 17,290,316 (1.43) | 1,794 / 6,249,994 (2.87) | 171 / 3,160,206 (0.54) | 516 / 7,880,116 (0.65) |
| 13 Dec-2 Jan | 4,901 / 33,869,084 (1.45) | 3,572 / 18,109,882 (1.97) | 915 / 10,943,494 (0.84) | 414 / 4,815,710 (0.86) |
| 3 Jan-23 Jan | 7,959 / 23,635,116 (3.37) | 5,864 / 12,863,158 (4.56) | 1,736 / 8,992,476 (1.93) | 359 / 1,779,482 (2.02) |
| 24 Jan-14 Feb | 1,444 / 3,544,088 (4.07) | 897 / 1,844,230 (4.86) | 503 / 1,588,412 (3.17) | 44 / 111,445 (3.95) |

**Supplementary Table 2.** Cases, deaths, followup days, and deaths per 10,000 days of followup, by SGTF and non-SGTF, cross-tabulated by date range, geographical region, age group, and IMD group. The number in parentheses is the rate of death per 10,000 days of follow-up. EE, LD, SE: East of England, London, South East (regions in which B.1.1.7 was first detected); ML, NEY, NW, SW: Midlands, North East & Yorkshire, North West, South West (other regions).

|  |  | 2020-11-01 - 2020-11-20 |  |
| --- | --- | --- | --- |
|  |  | EE, LD, SE | ML, NEY, NW, SW |
| Age 1 - 54 | IMD 1 - 2 | SGTF: 1086 cases, 0 deaths / 30408 days (0)<br>Non-SGTF: 4453 cases, 2 deaths / 124670 days (0.16) | SGTF: 942 cases, 0 deaths / 26376 days (0)<br>Non-SGTF: 44032 cases, 17 deaths / 1232673 days (0.14) |
|  | IMD 3 - 8 | SGTF: 4827 cases, 0 deaths / 135156 days (0)<br>Non-SGTF: 24448 cases, 5 deaths / 684449 days (0.07) | SGTF: 1659 cases, 0 deaths / 46452 days (0)<br>Non-SGTF: 68497 cases, 17 deaths / 1917679 days (0.09) |
|  | IMD 9 - 10 | SGTF: 1368 cases, 0 deaths / 38304 days (0)<br>Non-SGTF: 7618 cases, 1 deaths / 213286 days (0.05) | SGTF: 369 cases, 0 deaths / 10332 days (0)<br>Non-SGTF: 16586 cases, 4 deaths / 464346 days (0.09) |
| Age 55 - 69 | IMD 1 - 2 | SGTF: 192 cases, 3 deaths / 5331 days (5.63)<br>Non-SGTF: 599 cases, 7 deaths / 16672 days (4.2) | SGTF: 159 cases, 0 deaths / 4452 days (0)<br>Non-SGTF: 7885 cases, 49 deaths / 220189 days (2.23) |
|  | IMD 3 - 8 | SGTF: 800 cases, 4 deaths / 22353 days (1.79)<br>Non-SGTF: 3874 cases, 11 deaths / 108348 days (1.02) | SGTF: 354 cases, 0 deaths / 9912 days (0)<br>Non-SGTF: 15729 cases, 46 deaths / 439885 days (1.05) |
|  | IMD 9 - 10 | SGTF: 277 cases, 0 deaths / 7756 days (0)<br>Non-SGTF: 1563 cases, 0 deaths / 43764 days (0) | SGTF: 92 cases, 0 deaths / 2576 days (0)<br>Non-SGTF: 4280 cases, 8 deaths / 119765 days (0.67) |
| Age 70 - 84 | IMD 1 - 2 | SGTF: 47 cases, 1 deaths / 1296 days (7.72)<br>Non-SGTF: 124 cases, 2 deaths / 3452 days (5.79) | SGTF: 41 cases, 3 deaths / 1112 days (26.98)<br>Non-SGTF: 2195 cases, 97 deaths / 59987.5 days (16.17) |
|  | IMD 3 - 8 | SGTF: 166 cases, 4 deaths / 4591 days (8.71)<br>Non-SGTF: 1057 cases, 16 deaths / 29358 days (5.45) | SGTF: 117 cases, 3 deaths / 3226 days (9.3)<br>Non-SGTF: 4402 cases, 143 deaths / 121187 days (11.8) |
|  | IMD 9 - 10 | SGTF: 64 cases, 0 deaths / 1792 days (0)<br>Non-SGTF: 436 cases, 8 deaths / 12067 days (6.63) | SGTF: 26 cases, 1 deaths / 722 days (13.85)<br>Non-SGTF: 1225 cases, 38 deaths / 33783 days (11.25) |
| Age 85 - 120 | IMD 1 - 2 | SGTF: 3 cases, 1 deaths / 83 days (120.48)<br>Non-SGTF: 24 cases, 1 deaths / 658 days (15.2) | SGTF: 4 cases, 0 deaths / 112 days (0)<br>Non-SGTF: 360 cases, 61 deaths / 9140 days (66.74) |
|  | IMD 3 - 8 | SGTF: 11 cases, 3 deaths / 276 days (108.7)<br>Non-SGTF: 136 cases, 12 deaths / 3621 days (33.14) | SGTF: 16 cases, 1 deaths / 433 days (23.09)<br>Non-SGTF: 711 cases, 96 deaths / 18480.5 days (51.95) |
|  | IMD 9 - 10 | SGTF: 6 cases, 1 deaths / 157 days (63.69)<br>Non-SGTF: 76 cases, 13 deaths / 1956 days (66.46) | SGTF: 5 cases, 1 deaths / 125 days (80)<br>Non-SGTF: 211 cases, 29 deaths / 5455 days (53.16) |
|  |  | 2020-11-22 - 2020-12-11 |  |
|  |  | EE, LD, SE | ML, NEY, NW, SW |
| Age 1 - 54 | IMD 1 - 2 | SGTF: 3850 cases, 1 deaths / 107787 days (0.09)<br>Non-SGTF: 3119 cases, 2 deaths / 87300 days (0.23) | SGTF: 1470 cases, 0 deaths / 41160 days (0)<br>Non-SGTF: 21373 cases, 11 deaths / 598239.5 days (0.18) |
|  | IMD 3 - 8 | SGTF: 19213 cases, 10 deaths / 537848 days (0.19)<br>Non-SGTF: 15504 cases, 4 deaths / 434052 days (0.09) | SGTF: 2844 cases, 0 deaths / 79632 days (0)<br>Non-SGTF: 31351 cases, 5 deaths / 877736 days (0.06) |
|  | IMD 9 - 10 | SGTF: 5945 cases, 0 deaths / 166460 days (0)<br>Non-SGTF: 4113 cases, 0 deaths / 115164 days (0) | SGTF: 769 cases, 0 deaths / 21532 days (0)<br>Non-SGTF: 7831 cases, 2 deaths / 219241 days (0.09) |
| Age 55 - 69 | IMD 1 - 2 | SGTF: 474 cases, 3 deaths / 13237 days (2.27)<br>Non-SGTF: 379 cases, 3 deaths / 10591 days (2.83) | SGTF: 176 cases, 1 deaths / 4904 days (2.04)<br>Non-SGTF: 3060 cases, 35 deaths / 85105 days (4.11) |
|  | IMD 3 - 8 | SGTF: 2361 cases, 19 deaths / 65965 days (2.88)<br>Non-SGTF: 2023 cases, 9 deaths / 56548 days (1.59) | SGTF: 401 cases, 1 deaths / 11220 days (0.89)<br>Non-SGTF: 6093 cases, 28 deaths / 170247 days (1.64) |
|  | IMD 9 - 10 | SGTF: 821 cases, 2 deaths / 22972 days (0.87)<br>Non-SGTF: 689 cases, 2 deaths / 19261 days (1.04) | SGTF: 116 cases, 1 deaths / 3240 days (3.09)<br>Non-SGTF: 1608 cases, 12 deaths / 44909 days (2.67) |
| Age 70 - 84 | IMD 1 - 2 | SGTF: 75 cases, 5 deaths / 2054 days (24.34)<br>Non-SGTF: 72 cases, 4 deaths / 1926 days (20.77) | SGTF: 37 cases, 4 deaths / 997 days (40.12)<br>Non-SGTF: 821 cases, 37 deaths / 22445 days (16.48) |
|  | IMD 3 - 8 | SGTF: 437 cases, 20 deaths / 11963 days (16.72)<br>Non-SGTF: 483 cases, 12 deaths / 13356 days (8.98) | SGTF: 82 cases, 6 deaths / 2198 days (27.3)<br>Non-SGTF: 1689 cases, 67 deaths / 46243 days (14.49) |
|  | IMD 9 - 10 | SGTF: 152 cases, 7 deaths / 4144 days (16.89)<br>Non-SGTF: 155 cases, 8 deaths / 4181 days (19.13) | SGTF: 32 cases, 0 deaths / 896 days (0)<br>Non-SGTF: 412 cases, 16 deaths / 11275 days (14.19) |
| Age 85 - 120 | IMD 1 - 2 | SGTF: 29 cases, 10 deaths / 686 days (145.77)<br>Non-SGTF: 9 cases, 2 deaths / 218 days (91.74) | SGTF: 12 cases, 0 deaths / 336 days (0)<br>Non-SGTF: 206 cases, 32 deaths / 5233 days (61.15) |
|  | IMD 3 - 8 | SGTF: 68 cases, 8 deaths / 1788 days (44.74)<br>Non-SGTF: 86 cases, 8 deaths / 2251 days (35.54) | SGTF: 22 cases, 7 deaths / 476.5 days (146.9)<br>Non-SGTF: 411 cases, 63 deaths / 10512 days (59.93) |
|  | IMD 9 - 10 | SGTF: 28 cases, 9 deaths / 643 days (139.97)<br>Non-SGTF: 35 cases, 6 deaths / 898 days (66.82) | SGTF: 4 cases, 0 deaths / 112 days (0)<br>Non-SGTF: 116 cases, 14 deaths / 2972 days (47.11) |

**Supplementary Table 2, continued.**
|  |  | 2020-12-13 - 2021-01-01 |  |
| --- | --- | --- | --- |
|  |  | EE, LD, SE | ML, NEY, NW, SW |
| Age 1 - 54 | IMD 1 - 2 | SGTF: 14181 cases, 6 deaths / 397009 days (0.15)<br>Non-SGTF: 2467 cases, 2 deaths / 69033 days (0.29) | SGTF: 13313 cases, 16 deaths / 372557 days (0.43)<br>Non-SGTF: 15724 cases, 9 deaths / 440126 days (0.2) |
|  | IMD 3 - 8 | SGTF: 76713 cases, 33 deaths / 2147551 days (0.15)<br>Non-SGTF: 13440 cases, 2 deaths / 376279 days (0.05) | SGTF: 23994 cases, 16 deaths / 671660 days (0.24)<br>Non-SGTF: 25618 cases, 15 deaths / 717092 days (0.21) |
|  | IMD 9 - 10 | SGTF: 23648 cases, 9 deaths / 662060 days (0.14)<br>Non-SGTF: 3930 cases, 0 deaths / 110040 days (0) | SGTF: 7425 cases, 1 deaths / 207886 days (0.05)<br>Non-SGTF: 6527 cases, 0 deaths / 182756 days (0) |
| Age 55 - 69 | IMD 1 - 2 | SGTF: 2030 cases, 27 deaths / 56481 days (4.78)<br>Non-SGTF: 361 cases, 1 deaths / 10106 days (0.99) | SGTF: 1949 cases, 30 deaths / 54217 days (5.53)<br>Non-SGTF: 2649 cases, 24 deaths / 73884 days (3.25) |
|  | IMD 3 - 8 | SGTF: 12258 cases, 96 deaths / 342029 days (2.81)<br>Non-SGTF: 2177 cases, 10 deaths / 60854 days (1.64) | SGTF: 4926 cases, 39 deaths / 137545 days (2.84)<br>Non-SGTF: 5931 cases, 19 deaths / 165820 days (1.15) |
|  | IMD 9 - 10 | SGTF: 4717 cases, 23 deaths / 131754 days (1.75)<br>Non-SGTF: 846 cases, 1 deaths / 23680 days (0.42) | SGTF: 1692 cases, 6 deaths / 47331 days (1.27)<br>Non-SGTF: 1630 cases, 3 deaths / 45599 days (0.66) |
| Age 70 - 84 | IMD 1 - 2 | SGTF: 349 cases, 21 deaths / 9454 days (22.21)<br>Non-SGTF: 58 cases, 1 deaths / 1610 days (6.21) | SGTF: 443 cases, 28 deaths / 12013 days (23.31)<br>Non-SGTF: 708 cases, 40 deaths / 19256 days (20.77) |
|  | IMD 3 - 8 | SGTF: 2601 cases, 99 deaths / 71448 days (13.86)<br>Non-SGTF: 499 cases, 11 deaths / 13811 days (7.96) | SGTF: 1255 cases, 68 deaths / 34198 days (19.88)<br>Non-SGTF: 1722 cases, 68 deaths / 47165 days (14.42) |
|  | IMD 9 - 10 | SGTF: 1047 cases, 29 deaths / 28936 days (10.02)<br>Non-SGTF: 187 cases, 6 deaths / 5145 days (11.66) | SGTF: 449 cases, 19 deaths / 12344 days (15.39)<br>Non-SGTF: 437 cases, 11 deaths / 12104 days (9.09) |
| Age 85 - 120 | IMD 1 - 2 | SGTF: 52 cases, 12 deaths / 1298 days (92.45)<br>Non-SGTF: 3 cases, 2 deaths / 63 days (317.46) | SGTF: 63 cases, 10 deaths / 1607 days (62.23)<br>Non-SGTF: 145 cases, 21 deaths / 3721 days (56.44) |
|  | IMD 3 - 8 | SGTF: 284 cases, 39 deaths / 7322 days (53.26)<br>Non-SGTF: 65 cases, 6 deaths / 1696 days (35.38) | SGTF: 180 cases, 37 deaths / 4493 days (82.35)<br>Non-SGTF: 367 cases, 66 deaths / 9204 days (71.71) |
|  | IMD 9 - 10 | SGTF: 153 cases, 22 deaths / 3938.5 days (55.86)<br>Non-SGTF: 43 cases, 11 deaths / 987.5 days (111.39) | SGTF: 47 cases, 7 deaths / 1198 days (58.43)<br>Non-SGTF: 92 cases, 15 deaths / 2307 days (65.02) |
|  |  | 2021-01-03 - 2021-01-22 |  |
|  |  | EE, LD, SE | ML, NEY, NW, SW |
| Age 1 - 54 | IMD 1 - 2 | SGTF: 12871 cases, 6 deaths / 354017 days (0.17)<br>Non-SGTF: 696 cases, 1 deaths / 19256 days (0.52) | SGTF: 44708 cases, 48 deaths / 1222863 days (0.39)<br>Non-SGTF: 12412 cases, 5 deaths / 343288 days (0.15) |
|  | IMD 3 - 8 | SGTF: 61982 cases, 40 deaths / 1702497 days (0.23)<br>Non-SGTF: 3773 cases, 1 deaths / 104360 days (0.1) | SGTF: 62155 cases, 39 deaths / 1701476 days (0.23)<br>Non-SGTF: 17137 cases, 12 deaths / 473824 days (0.25) |
|  | IMD 9 - 10 | SGTF: 14006 cases, 10 deaths / 384876.5 days (0.26)<br>Non-SGTF: 941 cases, 1 deaths / 26011 days (0.38) | SGTF: 14185 cases, 8 deaths / 388859 days (0.21)<br>Non-SGTF: 3510 cases, 3 deaths / 97069 days (0.31) |
| Age 55 - 69 | IMD 1 - 2 | SGTF: 2024 cases, 18 deaths / 55339 days (3.25)<br>Non-SGTF: 113 cases, 2 deaths / 3105 days (6.44) | SGTF: 7758 cases, 129 deaths / 210715 days (6.12)<br>Non-SGTF: 2368 cases, 15 deaths / 65117 days (2.3) |
|  | IMD 3 - 8 | SGTF: 11460 cases, 79 deaths / 313889 days (2.52)<br>Non-SGTF: 646 cases, 4 deaths / 17831 days (2.24) | SGTF: 14056 cases, 123 deaths / 382714 days (3.21)<br>Non-SGTF: 4280 cases, 26 deaths / 117914 days (2.2) |
|  | IMD 9 - 10 | SGTF: 3464 cases, 23 deaths / 95017 days (2.42)<br>Non-SGTF: 218 cases, 0 deaths / 5997 days (0) | SGTF: 3787 cases, 22 deaths / 103572 days (2.12)<br>Non-SGTF: 982 cases, 7 deaths / 27089 days (2.58) |
| Age 70 - 84 | IMD 1 - 2 | SGTF: 387 cases, 11 deaths / 10546 days (10.43)<br>Non-SGTF: 29 cases, 2 deaths / 767 days (26.08) | SGTF: 1941 cases, 152 deaths / 50738 days (29.96)<br>Non-SGTF: 702 cases, 56 deaths / 18460.5 days (30.34) |
|  | IMD 3 - 8 | SGTF: 2246 cases, 95 deaths / 60410.5 days (15.73)<br>Non-SGTF: 156 cases, 1 deaths / 4279 days (2.34) | SGTF: 3779 cases, 242 deaths / 99990 days (24.2)<br>Non-SGTF: 1371 cases, 56 deaths / 36970.5 days (15.15) |
|  | IMD 9 - 10 | SGTF: 817 cases, 32 deaths / 21875 days (14.63)<br>Non-SGTF: 58 cases, 4 deaths / 1546 days (25.87) | SGTF: 1009 cases, 41 deaths / 27043 days (15.16)<br>Non-SGTF: 365 cases, 14 deaths / 9867 days (14.19) |
| Age 85 - 120 | IMD 1 - 2 | SGTF: 50 cases, 8 deaths / 1225 days (65.31)<br>Non-SGTF: 5 cases, 1 deaths / 111 days (90.09) | SGTF: 427 cases, 97 deaths / 10060 days (96.42)<br>Non-SGTF: 215 cases, 47 deaths / 5074 days (92.63) |
|  | IMD 3 - 8 | SGTF: 431 cases, 63 deaths / 10772 days (58.48)<br>Non-SGTF: 33 cases, 7 deaths / 792 days (88.38) | SGTF: 985 cases, 201 deaths / 23329.5 days (86.16)<br>Non-SGTF: 388 cases, 60 deaths / 9692 days (61.91) |
|  | IMD 9 - 10 | SGTF: 229 cases, 46 deaths / 5580 days (82.44)<br>Non-SGTF: 19 cases, 1 deaths / 512 days (19.53) | SGTF: 266 cases, 59 deaths / 6271 days (94.08)<br>Non-SGTF: 100 cases, 13 deaths / 2584 days (50.31) |
|  |  | 2021-01-24 - 2021-02-14 |  |
|  |  | EE, LD, SE | ML, NEY, NW, SW |
| Age 1 - 54 | IMD 1 - 2 | SGTF: 3888 cases, 2 deaths / 52209 days (0.38)<br>Non-SGTF: 114 cases, 0 deaths / 1656.5 days (0) | SGTF: 27972 cases, 13 deaths / 326729.5 days (0.4)<br>Non-SGTF: 1846 cases, 1 deaths / 26474.5 days (0.38) |
|  | IMD 3 - 8 | SGTF: 20931 cases, 4 deaths / 278769 days (0.14)<br>Non-SGTF: 584 cases, 0 deaths / 8290.5 days (0) | SGTF: 36411 cases, 9 deaths / 432872 days (0.21)<br>Non-SGTF: 2458 cases, 1 deaths / 35407 days (0.28) |
|  | IMD 9 - 10 | SGTF: 5353 cases, 0 deaths / 69844.5 days (0)<br>Non-SGTF: 161 cases, 0 deaths / 2380.5 days (0) | SGTF: 7623 cases, 1 deaths / 91848 days (0.11)<br>Non-SGTF: 498 cases, 0 deaths / 7288.5 days (0) |
| Age 55 - 69 | IMD 1 - 2 | SGTF: 590 cases, 5 deaths / 8061 days (6.2)<br>Non-SGTF: 17 cases, 0 deaths / 223 days (0) | SGTF: 4792 cases, 27 deaths / 56571 days (4.77)<br>Non-SGTF: 401 cases, 2 deaths / 5922.5 days (3.38) |
|  | IMD 3 - 8 | SGTF: 3753 cases, 15 deaths / 51132.5 days (2.93)<br>Non-SGTF: 113 cases, 2 deaths / 1683 days (11.88) | SGTF: 8245 cases, 45 deaths / 99120 days (4.54)<br>Non-SGTF: 655 cases, 3 deaths / 9578.5 days (3.13) |
|  | IMD 9 - 10 | SGTF: 1329 cases, 3 deaths / 17341.5 days (1.73)<br>Non-SGTF: 62 cases, 0 deaths / 891 days (0) | SGTF: 1993 cases, 2 deaths / 23714.5 days (0.84)<br>Non-SGTF: 172 cases, 0 deaths / 2462 days (0) |
| Age 70 - 84 | IMD 1 - 2 | SGTF: 109 cases, 1 deaths / 1557 days (6.42)<br>Non-SGTF: 0 cases, 0 deaths / 0 days (NaN) | SGTF: 1170 cases, 44 deaths / 14501 days (30.34)<br>Non-SGTF: 118 cases, 6 deaths / 1611 days (37.24) |
|  | IMD 3 - 8 | SGTF: 770 cases, 26 deaths / 10242.5 days (25.38)<br>Non-SGTF: 27 cases, 0 deaths / 415 days (0) | SGTF: 2006 cases, 82 deaths / 24108 days (34.01)<br>Non-SGTF: 256 cases, 6 deaths / 3509.5 days (17.1) |
|  | IMD 9 - 10 | SGTF: 313 cases, 11 deaths / 4040.5 days (27.22)<br>Non-SGTF: 10 cases, 0 deaths / 148 days (0) | SGTF: 477 cases, 19 deaths / 5939 days (31.99)<br>Non-SGTF: 58 cases, 2 deaths / 755.5 days (26.47) |
| Age 85 - 120 | IMD 1 - 2 | SGTF: 11 cases, 2 deaths / 167 days (119.76)<br>Non-SGTF: 0 cases, 0 deaths / 0 days (NaN) | SGTF: 357 cases, 42 deaths / 4102.5 days (102.38)<br>Non-SGTF: 54 cases, 2 deaths / 848 days (23.58) |
|  | IMD 3 - 8 | SGTF: 198 cases, 21 deaths / 2614 days (80.34)<br>Non-SGTF: 10 cases, 2 deaths / 114 days (175.44) | SGTF: 769 cases, 93 deaths / 9061.5 days (102.63)<br>Non-SGTF: 119 cases, 13 deaths / 1528.5 days (85.05) |
|  | IMD 9 - 10 | SGTF: 112 cases, 10 deaths / 1409 days (70.97)<br>Non-SGTF: 3 cases, 0 deaths / 41 days (0) | SGTF: 207 cases, 26 deaths / 2458 days (105.78)<br>Non-SGTF: 15 cases, 4 deaths / 217 days (184.33) |

## Supplementary Note 1

### Models with interaction terms

In the main text (section “Cox regression analyses”), we briefly describe results obtained from models including interactions between SGTF and other covariates, as well as interactions between other covariates and time since positive test, in our complete-cases analysis.

As stated in the main text, in our analysis of the effect of SGTF on the hazard of death due to COVID-19, we found no evidence that the effect of SGTF varied by age group, sex, IMD, ethnicity, or residence type. In an earlier analysis using data up to 25 January 2021, we did find a marginally significant interaction (likelihood ratio test 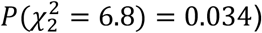 between SGTF and residence type indicating a higher hazard of death due to SGTF in care/nursing home residents, but this interaction is no longer present (likelihood ratio test 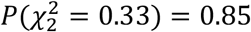) when analysing the more complete data set up to 14 February 2021.

We also describe in the main text a significant interaction between SGTF and time since positive test (henceforth, “time since positive test” is abbreviated as “time”, not to be confused with the date upon which the specimen was taken). In additional analyses, we also found significant interactions between age and time, sex and time, and place of residence and time.

Specifically, in our complete-cases analysis:

– the SGTF:time coefficient was estimated at 0.025 (standard error 0.0076), indicating a slightly longer time from specimen to death for individuals with SGTF (likelihood ratio test 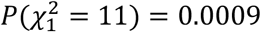);
– the age:time coefficient was estimated at –8.3×10^−4^ (SE 1.9×10^−4^), indicating a slightly shorter time from specimen to death for older individuals (likelihood ratio test 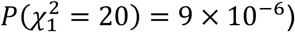;
– the sex_male_:time coefficient was estimated at 9.4×10^−3^ (SE 4.7×10^−3^), indicating a slightly longer time from specimen to death for males (likelihood ratio test 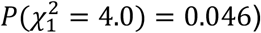; and
– the residence_care/nursing home_:time coefficient was estimated at –0.024 (SE 0.010), indicating a slightly shorter time from specimen to death for care home residents (likelihood ratio test 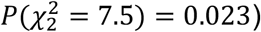.

By contrast, we did not find any significant interaction between ethnicity and time (likelihood ratio test 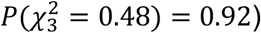 or between IMD decile and time (likelihood ratio test 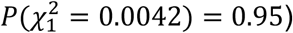.

## Supplementary Note 2

### CMMID COVID-19 Working Group acknowledgements

Funding statements for the CMMID COVID-19 working group are as follows. KvZ: KvZ is supported by the UK Foreign, Commonwealth and Development Office (FCDO)/Wellcome Trust Epidemic Preparedness Coronavirus research programme (ref. 221303/Z/20/Z), and Elrha’s Research for Health in Humanitarian Crises (R2HC) Programme, which aims to improve health outcomes by strengthening the evidence base for public health interventions in humanitarian crises. The R2HC programme is funded by the UK Government (FCDO), the Wellcome Trust, and the UK National Institute for Health Research (NIHR). SC: Wellcome Trust (grant: 208812/Z/17/Z). FYS: NIHR EPIC grant (16/137/109). SFunk: Wellcome Trust (grant: 210758/Z/18/Z), NIHR (NIHR200908). GFM: NTD Modelling Consortium by the Bill and Melinda Gates Foundation (OPP1184344). YJ: LSHTM, DHSC/UKRI COVID-19 Rapid Response Initiative. SRM: Wellcome Trust (grant: 210758/Z/18/Z). RL: Royal Society Dorothy Hodgkin Fellowship. WJE: European Commission (EpiPose 101003688), NIHR (NIHR200908). MQ: European Research Council Starting Grant (Action Number #757699); Bill and Melinda Gates Foundation (INV-001754). NRW: Medical Research Council (grant number MR/N013638/1). RME: HDR UK (grant: MR/S003975/1), MRC (grant: MC_PC 19065), NIHR (grant: NIHR200908). NGD: UKRI Research England; NIHR Health Protection Research Unit in Immunisation (NIHR200929); UK MRC (MC_PC_19065). JYL: Bill & Melinda Gates Foundation (INV-003174). MK: Foreign, Commonwealth and Development Office / Wellcome Trust. FK: Innovation Fund of the Joint Federal Committee (Grant number 01VSF18015), Wellcome Trust (UNS110424). DCT: No funding declared. JDM: Wellcome Trust (grant: 210758/Z/18/Z). AS: No funding declared. AMF: No funding declared. KP: Gates (INV-003174), European Commission (101003688). SFlasche: Wellcome Trust (grant: 208812/Z/17/Z). AJK: Wellcome Trust (grant: 206250/Z/17/Z), NIHR (NIHR200908). SA: Wellcome Trust (grant: 210758/Z/18/Z). BJQ: This research was partly funded by the National Institute for Health Research (NIHR) (16/137/109 & 16/136/46) using UK aid from the UK Government to support global health research. The views expressed in this publication are those of the author(s) and not necessarily those of the NIHR or the UK Department of Health and Social Care. BJQ is supported in part by a grant from the Bill and Melinda Gates Foundation (OPP1139859). TJ: RCUK/ESRC (grant: ES/P010873/1); UK PH RST; NIHR HPRU Modelling & Health Economics (NIHR200908). AR: NIHR (grant: PR-OD-1017-20002). GMK: UK Medical Research Council (grant: MR/P014658/1). MJ: Gates (INV-003174, INV-016832), NIHR (16/137/109, NIHR200929, NIHR200908), European Commission (EpiPose 101003688). YL: Gates (INV-003174), NIHR (16/137/109), European Commission (101003688). JW: NIHR Health Protection Research Unit and NIHR HTA. JH: Wellcome Trust (grant: 210758/Z/18/Z). KO’R: Bill and Melinda Gates Foundation (OPP1191821). YWDC: No funding declared. TWR: Wellcome Trust (grant: 206250/Z/17/Z). CIJ: Global Challenges Research Fund (GCRF) project ‘RECAP’ managed through RCUK and ESRC (ES/P010873/1). SRP: Bill and Melinda Gates Foundation (INV-016832). AE: The Nakajima Foundation. ESN: Gates (OPP1183986). NIB: Health Protection Research Unit (grant code NIHR200908). CJVA: European Research Council Starting Grant (Action number 757688). FGS: NIHR Health Protection Research Unit in Modelling & Health Economics, and in Immunisation. AG: European Commission (EpiPose 101003688). KA: Bill & Melinda Gates Foundation (OPP1157270, INV-016832). WW: MRC (grant MR/V027956/1). KEA: European Research Council Starting Grant (Action number 757688). RCB: European Commission (EpiPose 101003688). PK: This research was partly funded by the Royal Society under award RP\EA\180004, European Commission (101003688), Bill & Melinda Gates Foundation (INV-003174). HPG: This research was produced by CSIGN which is part of the EDCTP2 programme supported by the European Union (grant number RIA2020EF-2983-CSIGN). The views and opinions of authors expressed herein do not necessarily state or reflect those of EDCTP. This research is funded by the Department of Health and Social Care using UK Aid funding and is managed by the NIHR. The views expressed in this publication are those of the author(s) and not necessarily those of the Department of Health and Social Care (PR-OD-1017-20001). CABP: CABP is supported by the Bill & Melinda Gates Foundation (OPP1184344) and the UK Foreign, Commonwealth and Development Office (FCDO)/Wellcome Trust Epidemic Preparedness Coronavirus research programme (ref. 221303/Z/20/Z). OJB: Wellcome Trust (grant: 206471/Z/17/Z).

